# Mobility trends provide a leading indicator of changes in SARS-CoV-2 transmission

**DOI:** 10.1101/2020.05.07.20094441

**Authors:** Andrew C. Miller, Nicholas J. Foti, Joseph A. Lewnard, Nicholas P. Jewell, Carlos Guestrin, Emily B. Fox

## Abstract

Determining the impact of non-pharmaceutical interventions on transmission of the novel severe acute respiratory syndrome coronavirus 2 (SARS-CoV-2) is paramount for the design and deployment of effective public health policies. Incorporating Apple Maps mobility data into an epidemiological model of daily deaths and hospitalizations allowed us to estimate an explicit relationship between human mobility and transmission in the United States. We find that reduced mobility explains a large decrease in the effective reproductive number (*R_E_*) attained by April 1st and further identify state-to-state variation in the inferred transmission-mobility relationship. These findings indicate that simply relaxing stay-at-home orders can rapidly lead to outbreaks exceeding the scale of transmission that has occurred to date. Our findings provide quantitative guidance on the impact policies must achieve against transmission to safely relax social distancing measures.

---

As of April 25th, SARS-CoV-2 has caused approximately 2.9 million reported cases of coronavirus disease 2019 (COVID-19) and nearly 200,000 known deaths globally (*1*). At present, there are no specific preventative or therapeutic interventions, which will likely be the case for many months to come (*2*). As such, the public health response has centered on non-pharmaceutical interventions aiming to prevent transmission (*3*). While it is possible to contain transmission if infected individuals and their contacts are rapidly identified and isolated, short-comings in testing in many settings and the possibility of pre-symptomatic or asymptomatic transmission have necessitated broader social distancing measures to limit contact between susceptible and potentially infectious individuals (*4*). Such interventions have included closure of schools, businesses, and gathering places, and shelter-in-place orders. At present, an unprecedented proportion of the global population is living under such orders aiming to reduce contact and mobility (*5*). As of April 20th, over 316 million people in the U.S. in at least forty-two states have been urged to remain at home *(6*).

The economic, political, and mental health impacts of large-scale social distancing underscore the need to determine the efficacy of such interventions in reducing the burden of COVID-19. However, assessments of intervention effects are challenging to undertake: there is considerable delay between the start of an intervention and any reduction in cases and deaths that follows, and it may be unclear whether reductions owe to policy or other population-level behavior changes. The ubiquity of mobile phones make them a natural instrument for measuring movement in the population. Because real-time changes in contact patterns may impact infectious disease transmission, considerable interest has surrounded the possibility of using such data to assess the effect of social distancing interventions on COVID-19 burden *(7,8*). Previous studies have demonstrated that mobile phone data align with human migration patterns relevant to geographic spread of infectious diseases, including endemic diseases such as malaria and dengue *(9–11)* as well as COVID-19 *(12*). However, such data have not been used to study changes in contact impacting transmission at the scale of individual communities, or to assess the epidemiologic effects of behavioral interventions.

We aimed to assess the relationship between mobility trends and SARS-CoV-2 transmission using a mechanistic epidemiological model. We find that app-based mobility data provide a leading indicator of changes in transmission intensity, enabling real-time or forward-looking assessments of the effects of social-distancing measures on COVID-19 burden.

## Mobility trends data reveal differences in social mixing

While personal devices generate a wealth of information on human behavior, the use of these data for epidemiological purposes raises concerns about users’ privacy *(7*). In concordance with Apple’s strong stance on user privacy, the Apple Maps team has released a privacy-preserving data set that measures patterns in population mobility *(13*). The data reflect changes in volume of requests for directions in Apple Maps relative to the number of requests occurring on January 13, 2020. Data are not associated with user identifiers and are aggregated at the state level to preserve user privacy. We use this relative routing volume (RRV) data as a proxy for changes in person-to-person interactions in the population in our modeling framework.

Apple Maps mobility trends reveal interesting spatial and temporal patterns (Fig. 1). The traces are computed as the volume-weighted average over the three types of routes available — walking, transit, and drivin — expressed relative to a location-specific baseline (median volume from January 15 to February 15, 2020). The RRV data reveal a weekly cycle in population movement; Fridays and Saturdays in February showed about 25% higher volume than baseline, while volume as about 8% lower on Sundays. Holiday-associated changes in travel are also visible — there is a 39% increase in RRV for Valentine’s day (Feb. 14) across states, and the Saturday before (and day of) Mardis Gras festivities see RRV elevated to 88% (and 44%) of baseline within Louisiana.

**Figure 1:**
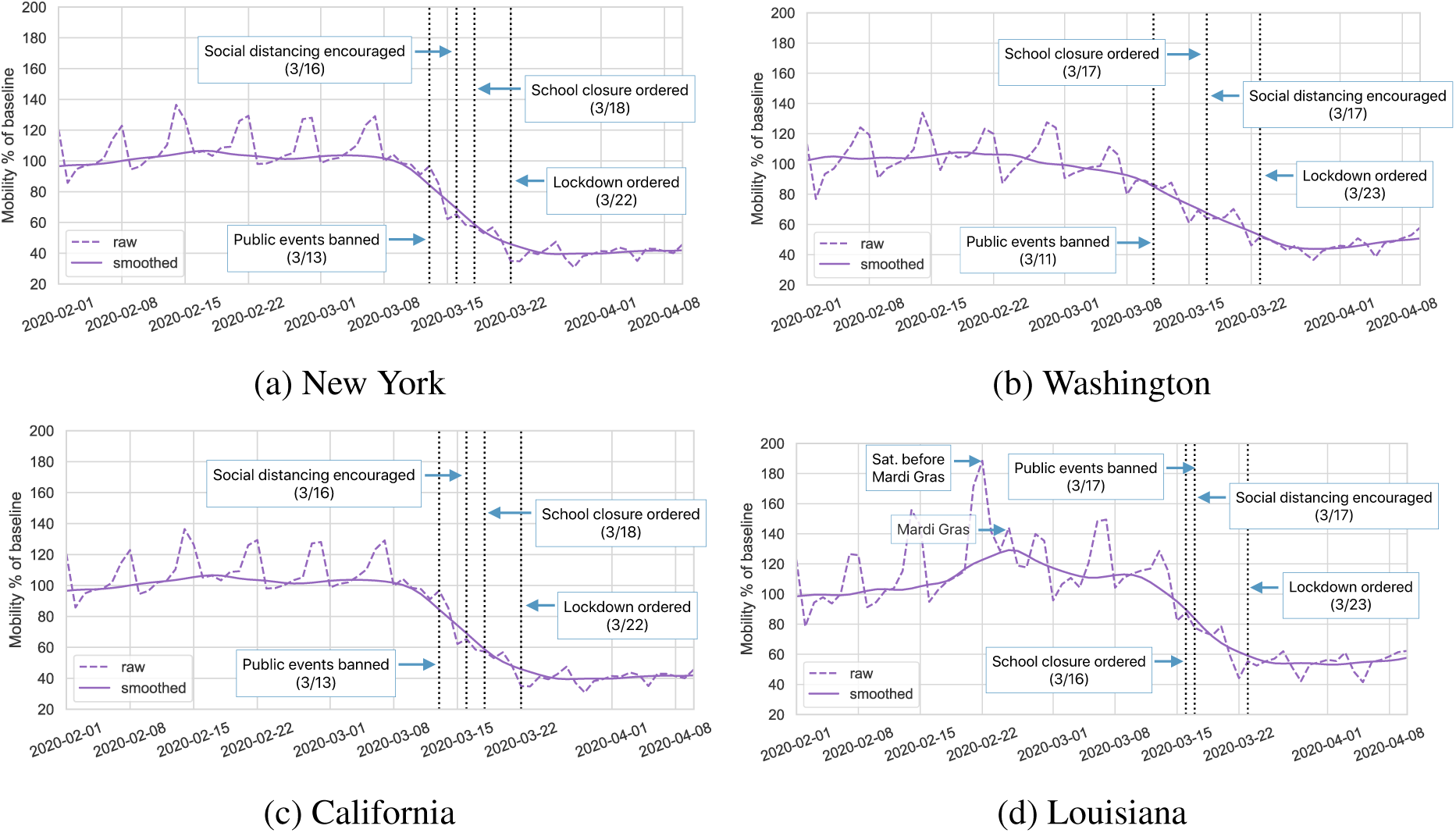
Mobility data and state-wide non-pharmaceutical interventions over time for New York, Washington, Florida, and Louisiana. Mobility data leads many interventions, particularly “lock down” or “shelter-in-place” orders and also reveals atypical patterns in activity.

The timing of implementation of non-pharmaceutical interventions against COVID-19 is associated with large drops in mobility, with reductions in RRV coinciding with — and in some cases preceding — statewide orders to shelter in place. Reduction in RRV before social distancing orders suggests that the population adopted risk-reducing behaviors as COVID-19 awareness spread. These mobility curves also reflect state-to-state variation in population response. In New York state, RRV fell to below 40% of January 13 baseline levels, while RRV in Louisiana remained above 50% of baseline. Capturing these phenomena without intensive surveys *(14*), or tracking of individuals’ specific locations, opens up new possibilities for non-invasive monitoring of behavior and mobility.

## Measuring transmission rates from mobility trends data

We next examined the hypothesis that changes in RRV measures predict changes in SARS-CoV-2 transmission in U.S. states. To this end, we develop an augmented epidemiologic model that describes the progression of the population through susceptible (*S*), exposed (*E*), infectious (*I*), hospitalized (*H*), and removed (*R*) compartments. The augmented model defines asymptomatic and symptomatic infectious paths. Symptomatic individuals can progress to one of two hospitalization compartments — one leading to recovery, the other to death — or to recovery/removal without hospitalization. Individuals acquire infection at rates proportional to the current prevalence of infection. We parameterize distributions over residency times in each compartment using estimates of the duration of each stage of the clinical progression of COVID-19 *(15-19)*. Probabilities of symptoms, hospitalization, and death are specified by separate parameters *(17,20)*. We extend the compartment model framework by incorporating mobility observations into a time-varying transmission parameter, *β_t_*. The model modulates a baseline (time-invariant) transmission parameter, *β*_0_, with a learned mobility function, *β_t_ = β*_0_ *· f* (Mobility*_t_*), where Mobility*_t_* denotes the fraction of baseline routing requests, as depicted in Fig. 1. Intuitively, *β*_0_ captures immutable aspects of transmission, e.g., the probability of infection upon contact between susceptible and infectious persons. The mobility function captures the reduction in transmissibility due to reduction in population mobility. We define this relationship, *f_λ_*(·), to be piece-wise linear and monotonic; we estimate λ from data.

The resulting model enables us to express reproductive numbers in terms of RRV data. The *basic reproductive number* is a fundamental quantity that describes the number of secondary cases generated by a single case in an entirely susceptible population, denoted *R_0_ = β*_0_ · τ_i_ where *τ_I_* is the average time spent infectious (in days). We define the *time-varying basic reproductive number R*_0,_*_t_* = *R*_0_ · *f*_λ_(Mobility*_t_*) to account for changes in *R*_0_ due to population behavior changing over time (i.e., mobility patterns). Another quantity of interest is the effective reproductive number *R_E_,_t_*, which accounts for the current state of the infection in the population, *R_E_,_t_ = R_0_,_t_ · (S_t_/N*) where *S_t_/N* is the proportion of individuals who remain susceptible at time t, and N is the population size.

We fit this model to counts of daily deaths (and daily hospitalizations where available) from two public data sources *(21,22*), including observations up to April 25th. We focus our analysis on eight states — New York, California, Washington, Texas, Georgia, Louisiana, Florida, and Massachusetts — selected to balance urban-rural variation and enough observed deaths (and hospitalizations) to fit the model. We adopted a Bayesian framework and computed the posterior distribution over all compartment times, rates, and mobility function parameters using a Markov Chain Monte Carlo sampler (*23*). For parameters without strong prior evidence, we used diffuse priors to propagate uncertainty into parameter estimates and forecasts. For each state, we compare models across prior informativeness, infection start date, and mobility function regularization strength, and present the model with the best held out log-likelihood (on eight-day forecasts; see SM for more details).

## Analysis of estimated reproductive numbers and mobility trends

Our analysis reveals differences across US states in pre- and post-intervention transmission as measured by *R_E_,_t_* in late February and March (Fig. 2). Since deaths lag infection by about three weeks, we depict estimated *R_E_,_t_* values up to April 1st. We found Louisiana and New York to have the highest pre-intervention transmission potential, with baseline *R*_0_ = 5.9 (3.5-9.5 95% CI) and 5.2 (2.8-8.9 95% CI) respectively. Florida and Texas had smaller pre-intervention *R*_0_ values, at around 3. This high value in *R*_0_ for New York comports with the observed intensity and explains observations of substantial cumulative infection prevalence based on population serosurveys (*24*). We plot state-to-state variation in baseline R_0_ estimates in SM Fig. 11. Among these states, we also find that only New York, Washington, and Louisiana had achieved *R_E_,_t_* < 1 (a threshold at which incidence rates can be expected to decline) by April 1st. Texas, Georgia, and Florida appeared to straddle *R_E_,_t_* = 1 at this time, while California and Massachusetts both retained *R_E_,_t_* > 1 on April 1st. These findings reflect observed increases in the daily death count continuing through the first three weeks of April in these states.

**Figure 2:**
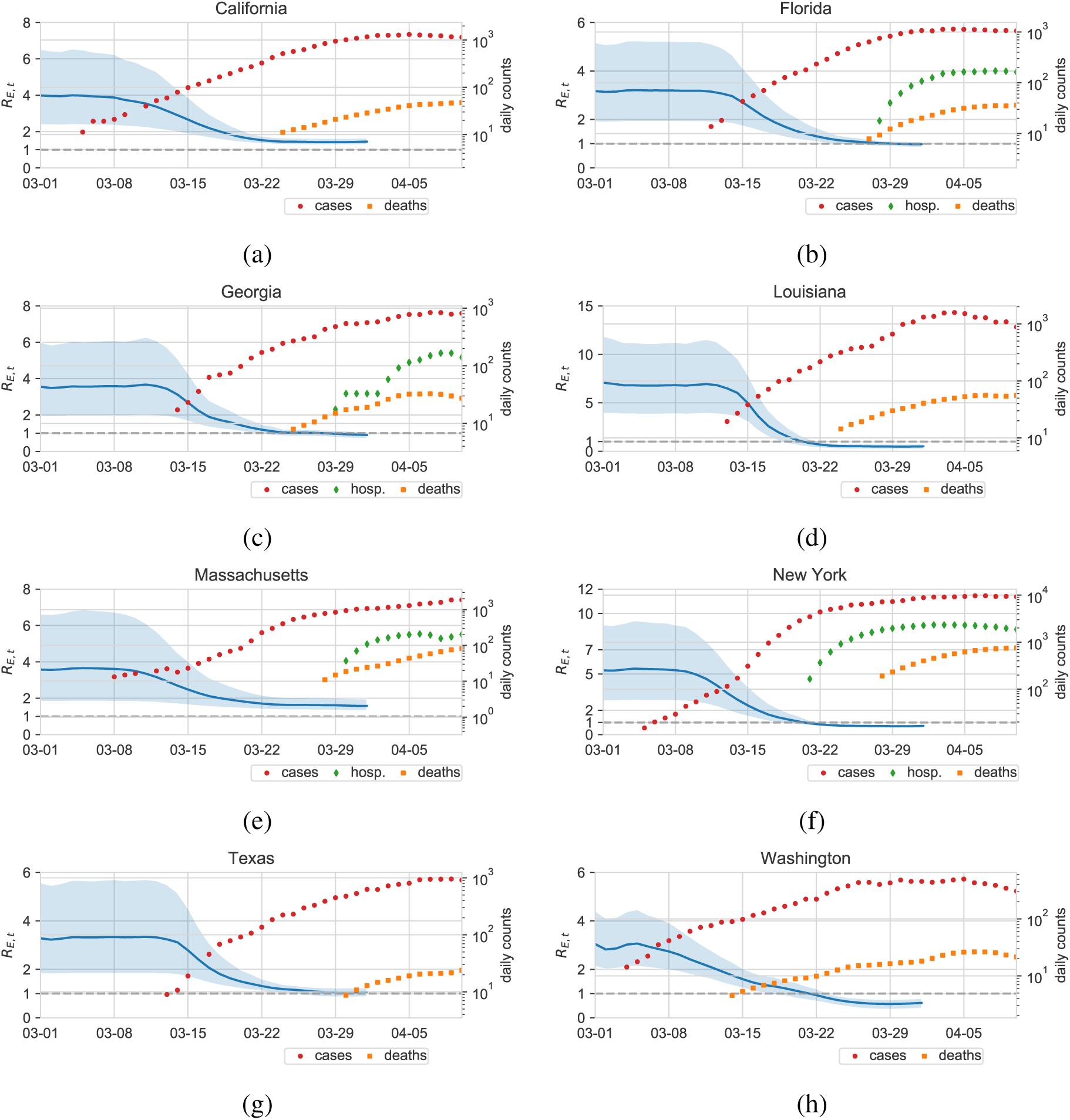
Inferred effective reproduction number *R_E,t_* for eight states, posterior mean and 95% credible intervals shown. Overlaid are daily case, hospitalization, and death counts (log scale, smoothed with a 7-day centered average). The effective reproduction number is modulated by time-varying mobility data and state-specific multiplier function.

The association between estimated *R_E_,_t_* and reduction in RRV varies in each state (Fig. 3). The mobility-driven factor, *R*_0_,*_t_* = *R*_0_ · *f* (Mobility_t_), falls below one at different levels of RRV depending on the state. For Louisiana, *R*_0_,*_t_* is reduced to one when RRV falls to 65% (58-75%) of baseline levels. Alternatively, New York’s *R*_0_,*_t_* falls below one when RRV is reduced to 48% (43-56%) of baseline. We also identified differences in the shape of relationships between RRV and *R*_0_,*_t_* by state. Reductions in RRV below 80% of baseline delivered diminishing returns in reducing *R*_0_,*_t_* in Louisiana, while the slope in New York was maximized at RRV around 50% of baseline.

**Figure 3:**
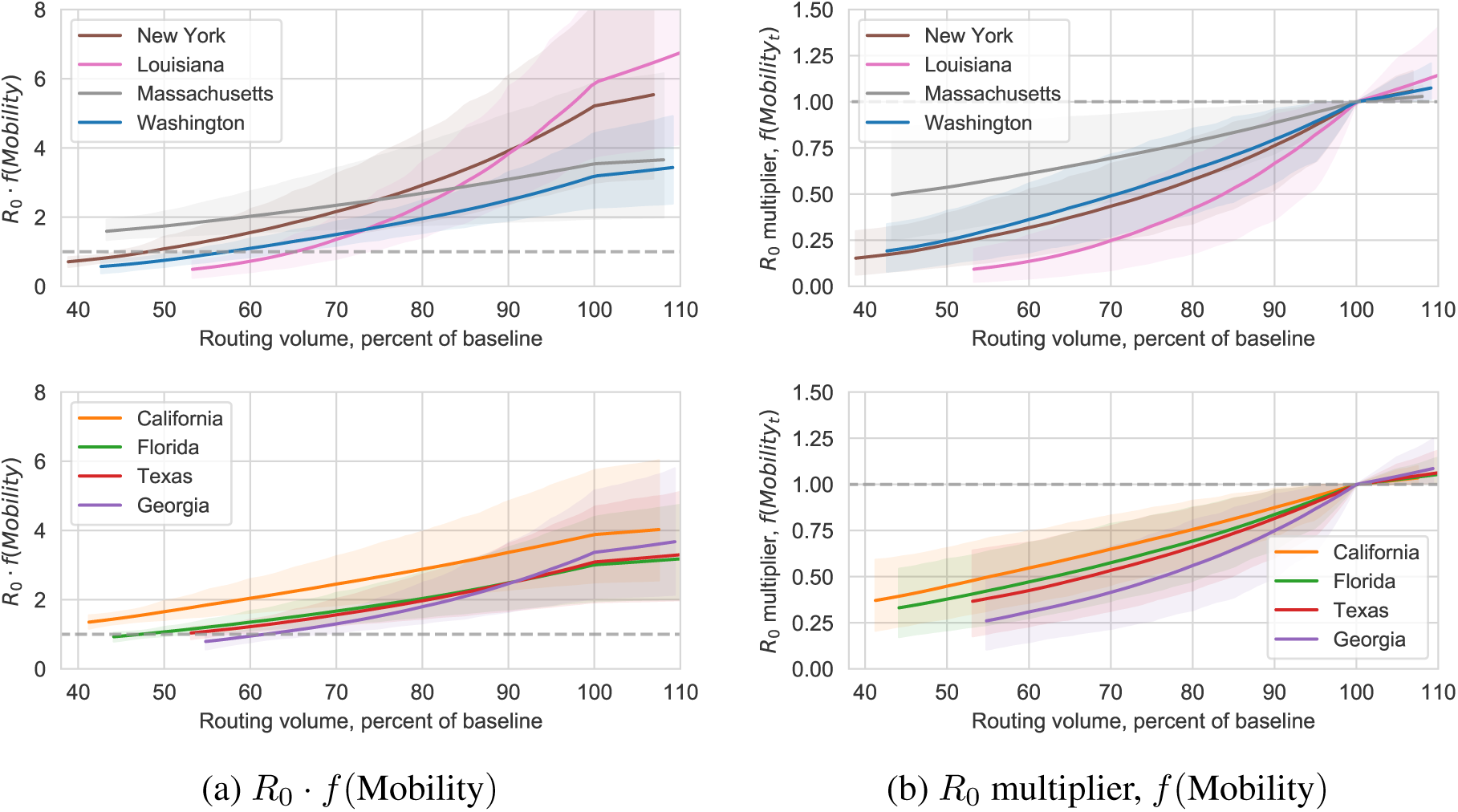
Left: Inferred relationship between reproduction number *R*_0,_*_t_* and mobility volume change, Mobility*_t_*, for eight states (split into two groups for clarity). The mobility reduction that corresponds to *R*_0,_*_t_* <= 1 varies from state to state. For Louisiana and Georgia, the value of *R*_0,_*_t_* falls below one when mobility reaches 60-70% of baseline. In contrast, *R*_0,_*_t_* for New York and California does not fall below (or approach) one until routing volume reaches 50% of baseline levels. Right: The *R*_0_ multiplier effect as a function of mobility. This relationship isolates the relative effect of mobility reduction on baseline *R*0.

To validate the estimated relationship between measured mobility and transmission, we generate an estimate of *R_E_,_t_* based solely on reported case data using the method of Wallinga and Teunis (*25*). We compared this alternative estimate of *R_E_,_t_* to the mobility data, identifying a significant positive relationship for each state (SM Fig. 13-17). The similar relationships found using two methods on disjoint sets of data provides evidence that changes in RRV can explain changes in *R_E,t_*. This finding substantiates the utility of mobility data as a real-time indicator of changes in transmission intensity.

## Evaluating policies to relax social distancing

Policy-makers currently face crucial questions about when and how existing social distancing policies can be relaxed without incurring large resurgences in transmission. We evaluated two potential policies for lifting social distancing measures to address the potential impacts on trajectories of COVID-19 burden.

We first consider relaxation of social distancing measures beginning June 1, 2020, allowing mobility volume to recover 10%, 25%, 50%, or 100% of the difference between levels observed as of April 1, 2020 and those observed before interventions came into effect. Fig. 4 depicts our forecasts of cumulative deaths, hospital occupancy, and the proportion of the population remaining susceptible through December 31, 2020. In all states analyzed, increasing mobility without other changes to behavior is expected to lead to substantially worse outcomes, in size or duration of the outbreak, than maintaining current mobility (see SM for results from additional states). In Florida, increasing mobility even a small amount results in *R_E,t_* > 1 inducing a massive secondary outbreak. The timing and extent of the outbreak is determined by the increase in mobility, with larger increases resulting in an earlier outbreak with cases concentrated over a shorter period of time. In Washington and Louisiana, potentially massive secondary outbreaks are only observed for lifting 50% or 100%, suggesting that there is more flexibility for Washington to allow increased mobility without additional changes to behavior. Since our model estimates that California has not yet achieved *R_E,t_* < 1 statewide, even maintaining mobility at the reduced level is expected to allow COVID-19 to continue spreading.

**Figure 4:**
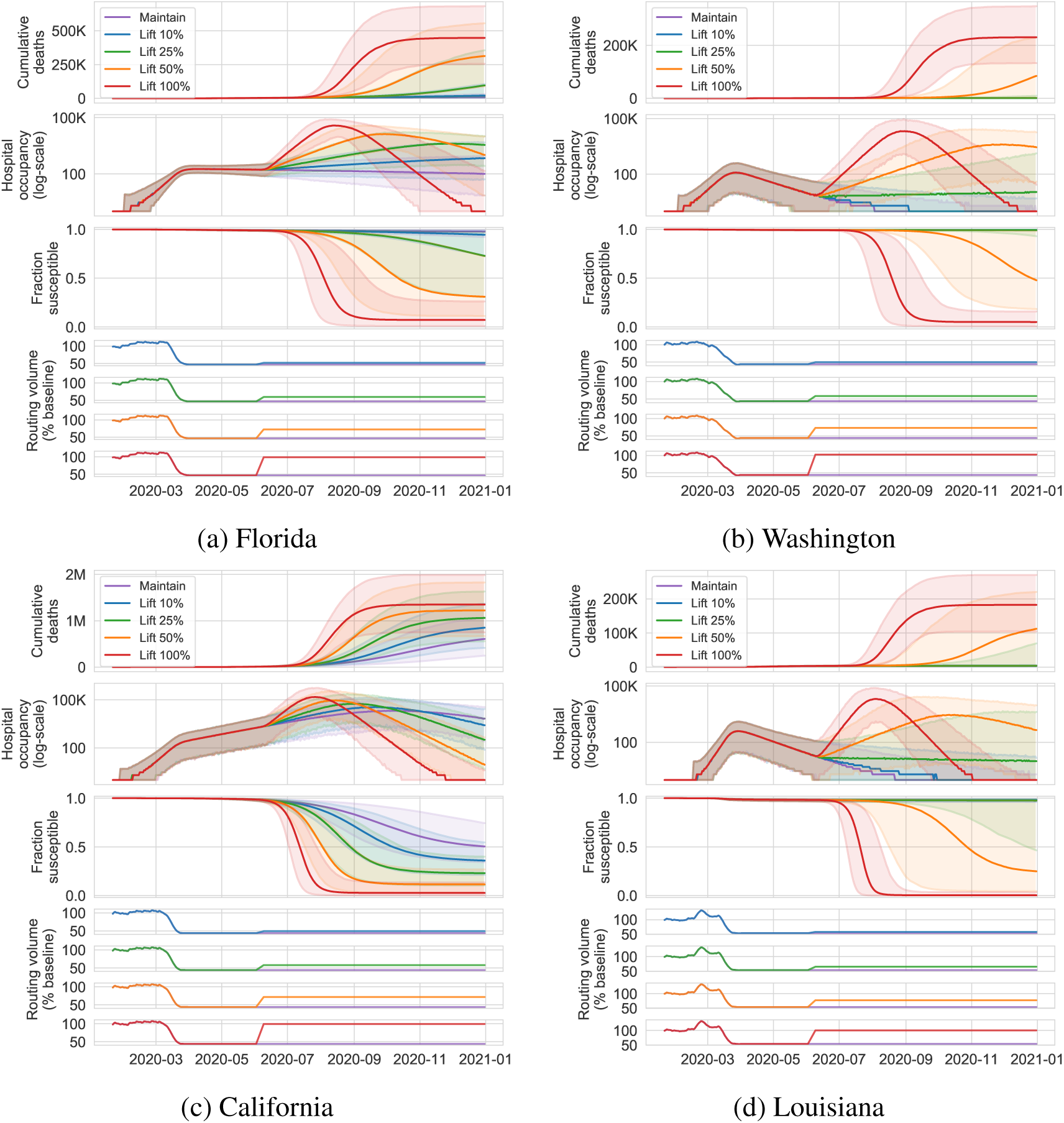
Median projections and 95% predictive intervals of cumulative deaths, hospital occupancy (log-scale), and fraction-susceptible through December 31st, 2020 when allowing mobility to increase 10%, 25%, 50%, and 100% of the way back to baseline. Plots zoomed in on the median projections.

As an alternative, we illustrate effects on transmission of a policy periodically lifting, beginning June 1st, for one week at a time, at 10-25% increases in RRV (Fig. 5). In all states we see that these periodic policies result in lower forecasted cumulative deaths and hospital occupancy than the previous policy. However, even forcing mobility to April 1st levels for four weeks, following one week of relaxation, may be inadequate to prevent resurgences in California and Florida exceeding pre-April burden. Lower baseline *R*_0_ values for Washington, and lower predicted prevalence of SARS-CoV-2 infection as of June 1, 2020, reduce the likelihood of a large-scale outbreak occurring in this state.

**Figure 5:**
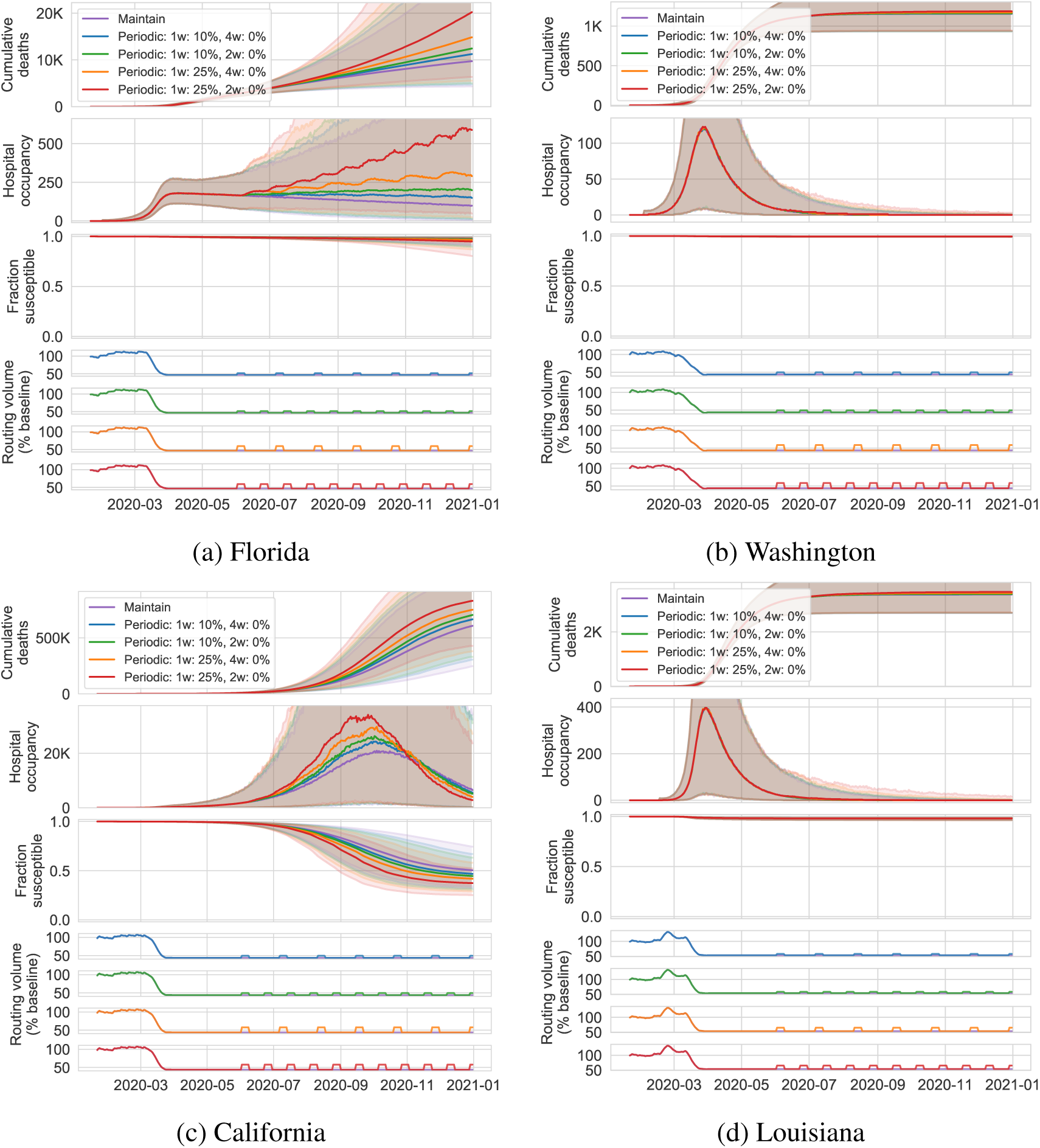
Median projections and 95% predictive intervals of cumulative deaths, hospital occupancy, and fraction-susceptible through December 31st, 2020 under the periodic policy that allows mobility to increase 10% or 25% of the way back to baseline for one week and then reimposing the April 1st mobility level for two or four weeks. Plots zoomed in on the median projections.

For mobility to safely increase within a state, reduction in the baseline transmission (i.e., *R*_0_) needs to be achieved in other ways e.g., by wearing masks or maintaining spatial distance in public. Fig. 6 reports for a number of states the percent reduction in *R*_0_ required to achieve *R*_0_,*_t_* = *R*_0_ · *f* (Mobility*_t_*) = 1 if mobility were to increase to 50% of baseline levels. This value reflects how the relationship in Fig. 3 must be altered through policies that reduce transmission for population mobility to increase toward pre-intervention levels while preserving epidemic control.

**Figure 6:**
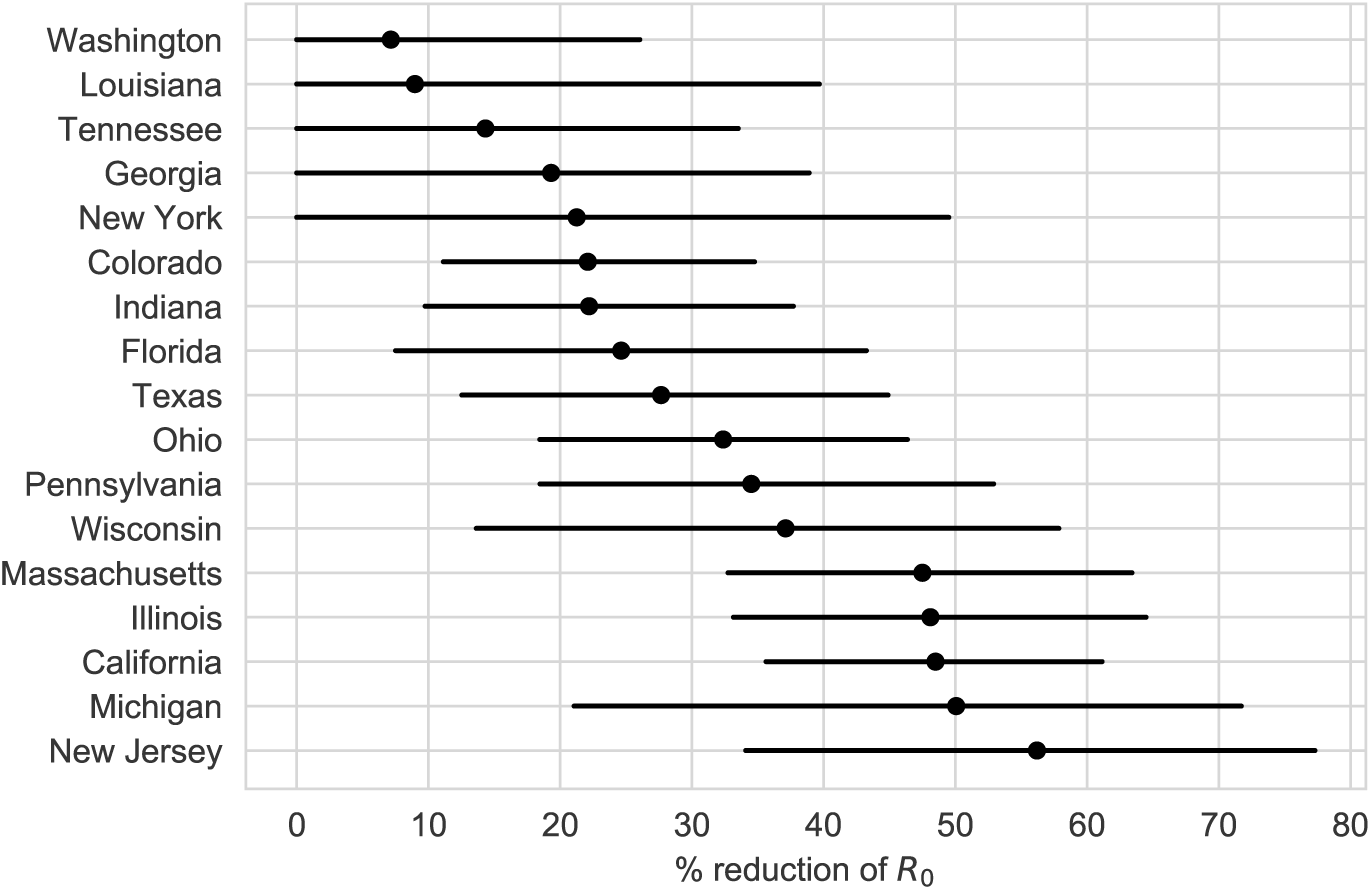
Percent reduction of mobility-independent R0 required to achieve *R_0,t_* ≤ 1 when mobility is 25% back to pre-intervention levels (relative to April 1, 2020). Reduction in baseline *R*_0_ would need to be achieved through a non-mobility change in behavior such as mask wearing, maintaining distance in public, and reducing the frequency of person-to-person interaction.

## Discussion

Examining the relationship between population-level mobility data and transmission rates, we found a strong relationship between mobility — as measured by Apple Maps routing volume — and transmission that varies from state to state. Changes in population behavior preceded implementation of statewide stay-at-home orders, and these data can allow us to attribute changes in transmission directly to this behavior change. Our findings suggest that mobility data can be used as a leading indicator of changes in transmission rates, supporting the evaluation of public health interventions. Our counterfactual analysis suggests that relaxing social distancing measures and returning to pre-intervention mobility *and behavior* could rapidly lead to transmission exceeding what the US has experienced to date. Going forward, the relationship between *R_E,t_* and population mobility must be altered going forward to avoid re-igniting the outbreak if population mobility is allowed to increase.

The modeling framework we present is extensible. Apple Maps mobility data is one measure of inter-personal interaction; other proxies can be incorporated into this approach as they become available (*7*, *8*). Unbiased infection data — collected through enhanced disease surveillance (*26*) or population serology studies — may allow more refined assessments of the relationship between population behavior and transmission.

Our analysis has limitations. Apple Maps data do have inherent biases. First, the demographics of Apple Maps users is unlikely to match the general population. Second, route requests are not a perfect indicator for movement or physical interaction and so are just a proxy. We also emphasize that the inferred relationship between population mobility and transmission reflects observations of the pandemic ramping up in the U.S. This relationship can change over time, however, through behavioral changes not strictly related to population mobility. For example, if mask-wearing and social-distancing continue, the baseline *β*_0_ (and thus *R*_0_) may be smaller than the pre-intervention *β*_0_. Indeed, our simulations show that we must reduce baseline transmission rates with non-mobility behavioral changes to return to even a fraction of pre-pandemic mobility patterns. Our model is a simplified description of population behavior. We do not address realistic clustered patterns of interaction of the population. Further, we do not model the effect of weather or temperature, which may contribute to changes in transmission intensity across seasons (*27*). Recent reports suggest that the overall death count is severely under-reported (*28*), which may contribute to bias in inferences of epidemic dynamics if reporting completeness varies over time (*29*).

Nonetheless, this work underscores the importance of new data sources for monitoring SARS-CoV-2 transmission. Privacy-preserving aggregated data from mobile phones can be used as a leading indicator of transmission rate changes. As states begin to relax social distancing measures, it is crucial to understand — and prepare for — the effects of increased population movement on disease transmission.

## Acknowledgments

We would like to acknowledge Alex Braunstein, Alyssa Glass Owara, Erin Gong, and Xiyan Wang of the Apple Maps team for providing assistance with the Mobility Trends data.

## Funding

JAL was supported by a grant from the University of California, Berkeley Population Center.

## Authors Contributions

ACM and NJF developed the model, conducted the analysis, and wrote the manuscript. JAL wrote and edited the manuscript. NPJ edited the manuscript. CG and EBF wrote and edited the manuscript.

## Competing Interests

ACM, NJF, CG, and EBF do not have any competing interests. JAL and NPJ have received honoraria from Kaiser Permanente unrelated to the current submission. JAL has received research grants and honoraria from Pfizer, research grants and honoraria from Merck, Sharp & Dohme, and honoraria from SutroVax unrelated to the current submission.

## Data Availability

The data used in the manuscript will be publicly available. Code to reproduce the results will be released on Github.

https://www.nytimes.com/article/coronavirus-county-data-us.html

https://covidtracking.com

https://www.apple.com/covid19/mobility

## Supplementary Materials

### Model Description

We developed an augmented version of the SEIR compartmental model that includes hospitalization, fatality, symptomatic, and asymptomatic infectious designations. We define the transmission parameter to vary in time as a function of population mobility measured as Apple maps routing volume. Through this learned relationship between transmission and mobility, we estimate (or simulate) the effect of mobility-based non-pharmaceutical interventions in different locations.

Infection spread in each state is described by the model depicted in Fig. 7. The compartments model different slices of the population at any given point in time

**Figure 7:**
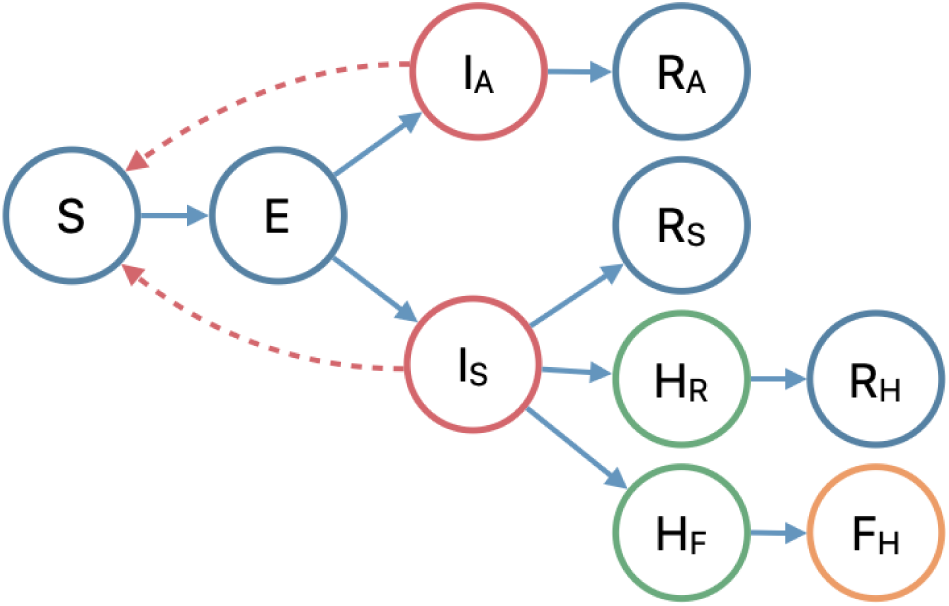
Augmented SEIR compartment model.

- *S*: susceptible population,
- *E*: exposed but non-infectious population,
- *I_S_*: eventually symptomatic and infectious population,
- *I_A_*: always asymptomatic and infectious population,
- *H_R_:* hospitalized cases that recover,
- *H_F_*: hospitalized cases that do not recover,
- *f_h_*: hospital fatality, and
- *R*: removed population.

The driving force behind the infection is the transmission of the virus from the infectious population, *I_S_* and *I_A_*, to the susceptible population, *S* (depicted in dotted arrows).

The dynamics of the model are parameterized by average compartment duration (i.e., dwell times). Each of the non-terminal compartments depends on a mean parameter that describes the average time within each compartment — e.g., the average time spent in the exposed state or the average time spent in the hospitalized state. Each non-terminal compartment is implemented with multiple stages to reflect a more realistic duration distribution, Erlang. We use two stages to represent each non-terminal compartment. Additionally, each branching component requires a rate parameter that divides the population flow between branches. These include the symptomatic rate (to *I_A_* or *I_S_*), the symptomatic hospitalization rate (to *H_R_, H_F_* or *R*), and the hospitalized fatality rate (to *F* or *R*). The compartment model parameters are defined as follows (time units are days):

- *τ_E_*: average dwell time in compartment E (incubation period),
- 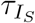: average dwell time in compartment I_s_ (infectious period),
- 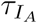: average dwell time in compartment I_A_ (asymptomatic infectious period),
- 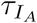: average hospital time for fatal cases,
- 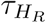: average hospital time for recovered cases,
- *η*: fraction of cases that are symptomatic,
- ν: fraction of symptomatic cases that are hospitalized,
- *μ*: fraction of hospitalized cases that are fatal.

Leveraging the evidence that patients can be asymptomatic and positive for weeks (*19*), we fix the durations of the asymptomatic and symptomatic cohorts to be equal, 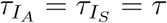1.

The dynamics between compartments are defined as:

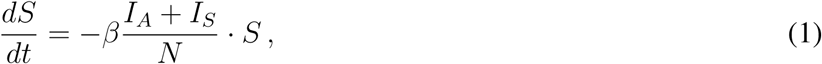

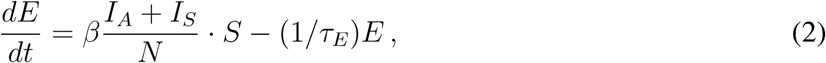

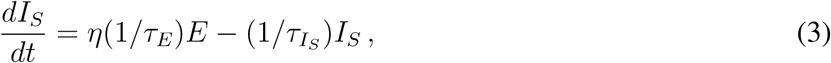

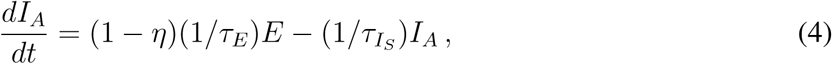

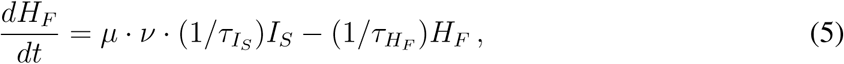

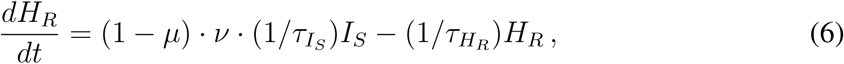

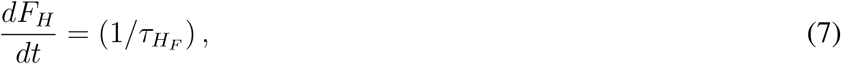

where *β* is a transmission parameter that relates to the basic reproductive number *R*_0_:

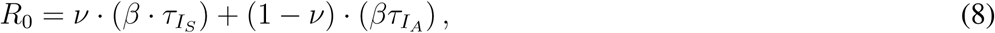

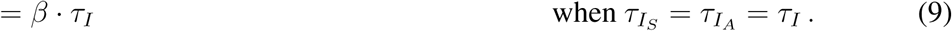

We directly parameterize the baseline *R*_0_ value, exploiting the above relationship between infectious duration, τ_I_, transmission parameter, *β*, and the basic reproductive number.

#### Mobility-driven transmission model

We allow the parameter *β* to vary in time, driven by observed mobility patterns. Denoting mobility volume change on day *t* as Mobility*_t_*, we define a time-varying *β* as

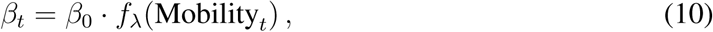

where *f_λ_*(·) is a piece-wise linear function with knots specified by parameters *λ*. The piece-wise linear function is monotonic by construction, and intersects 1 when Mobility*_t_* = 100%—when mobility is at baseline, *R*_0_;*t* is the baseline *R*_0_ = *β*_0_*τ*_I_.

#### Likelihood

We use a negative binomial likelihood for counts of observed daily deaths and hospitalizations. We found that it is crucial to incorporate an over-dispersion parameter for these counts, as variation in reporting can create larger observation variance than can be explained by a Poisson observation model. We place an exponential prior on the inverse over-dispersion parameter, *ϕ*, defining a preference for smaller observation variance in our prior beliefs. We fit separate *ϕ* = (*ϕ*^(^*^h^*^)^, *ϕ*^(^*^d^*^)^) values for hospitalization and death observations.

#### Prior specification

We place informative priors over the compartment dwell time and rate parameters culled from the recent COVID-19 literature.

For the rate of symptomatic cases, *η*, we adopt evidence from (*20*) that observes 43.2% (95% CI 32.2-54.7%) of confirmed SARS-CoV-2 infections were asymptomatic. Our symptomatic rate prior mean is 1 − .432 = .568 with a prior 95% credible interval that covers:25 and:75.

For the average hospitalization rate, *ν*, we use the age-specific estimates derived from travelers from Wuhan (*15*). Data from (*15*) are used to determine appropriate beta distribution for each ten-year age bucket. For each state, we average these age-specific hospitalization distributions over the state-specific population age distribution, approximating this weighted mixture with a beta distribution by matching 2.5 and 97.5 percentiles.

We use the hospital fatality rate estimate for California from (*17*) as an informative prior over *μ*, 17:8% (95% CI 14.3-22.2%), allowing for state-to-state variability.

Following (*16*), we place a truncated normal prior on *R*_0_ centered on 2.4. We test different strengths of the informativeness of this prior by setting different standard deviations. For the ‘weak’ version, we set the *R*_0_ scale to be 9, corresponding to an upper 97.5 percentile equal to *R*_0_ = 20:9. For the ‘medium’ version, we set the prior *R*_0_ scale to 6, corresponding to an upper 97.5 percentile equal to *R*_0_ = 14:4. For the ‘strong’ version, the prior *R*_0_ scale is set to 3, corresponding to an upper 97.5 percentile of *R*_0_ = 8:3. Note that placing a prior on *R*_0_ and a prior on *τ*_I_ induces a prior over *β*_0_.

For average time spent infectious, we see a wide range of evidence in the literature. (*18*) analyze Chinese cities using a dynamical model and infer that the average time spent infectious is 3.47 days (3.15, 3.73 95% credible interval). (*19*) examine virologic characteristics of a small sample (*n* = 12) of patients in the United States and find that average time between first and final positive rRT-PCR test is 16 days (12-21 95% CI). While this does not imply the average time spent infectious is 16 days, it suggests the use of a more diffuse prior on infectious time, 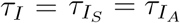. We place a Gamma prior distribution on *τ_I_* where the middle 95% of its mass covers the range 3 to 21.

For the incubation period, *τ_e_*, we adopt the mean inferred in (*18*), 3.69 days (3.30 - 3.96 95% CI) as our prior mean with a Gamma distribution. We inflate the prior variance to 1.

For the two hospitalized compartment times, we use evidence from (*17*). As we do not incorporate hospital recovery data, the recovery compartment time is only identified through the prior. As such, we use the average time from (*3*) of 10.7 days for 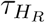 with a standard deviation of 1 day. For hospital fatality time, we again use evidence from (*17*) and set the prior mean of 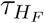 to be 13.7 days, with a standard deviation of 1.5 days.

We incorporate uncertainty into our initial conditions by placing a log-normal prior over *E*_0_*, I_A_*,_0_ and *I_S_*,_0_ with zero mean and standard deviation of two. Additionally, we search over the date for *t* = 0 by fitting separate models for *t*_0_ *=* {4, 5, 6} weeks before the first recorded COVID-19 death in each state.

Denote all unknown parameters as 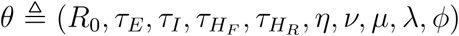, where *λ* parameterizes the mobility-transmission relationship *f_λ_(·)* and *ϕ* is the over-dispersion parameter of the negative-binomial likelihood. Note that we parameterize the model directly with *R*_0_ and derive *β*_0_ from *R*_0_ and *τ_i_*. Given the prior, likelihood, and structure of the differential equations, the full statistical model is

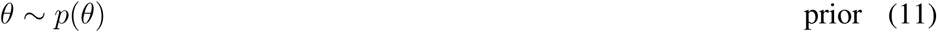

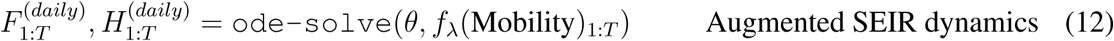

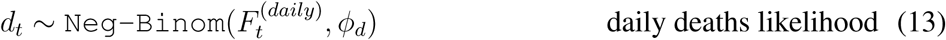

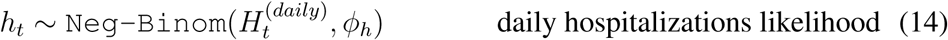

for observed daily deaths and hospitalization data *d*_1_*_:T_* and *h*_1_*_:T_* (where available), and 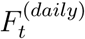 and 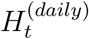 are the increase in *F_H_* and *h_r_* + *H_F_* compartments from day *t* − 1 to *t*, following (*30*).

## Model Fitting and Validation

### Inference

We use Bayesian inference to infer model parameters *θ* from daily observations *d*_1:_*_T_* and *h*_1_*_:T_* (note, some observations may be missing). We use the No U-turn Sampler (NUTS) (*23*), a variant of Hamiltonian Monte Carlo, to draw posterior samples for each model considered. We use a period of 1,000 samples for warm-up and step-size adaptation, and then draw 4 chains for 1,000 samples each. We use the NumPyro implementation of NUTS (*31*) and implement our models within the Python auto-differentiation framework JAX (*32*). For each model run, we diagnose convergence by inspecting the *ȓ* statistic for each sampled variable using the implementation in (*31*).

We focus on eight states — New York, California, Washington, Texas, Georgia, Louisiana, Florida, and Massachusetts — selected to balance urban-rural variation and enough observed deaths (and hospitalizations) to fit the model. Fig. 11 depicts the learned parameters and 95% credible intervals for the eight states considered in the main paper using the ‘medium’ prior.

### Validation forecasts and in sample fit

We validate our model by looking at statistics of eight-day-ahead and average eight-day-ahead forecasts. Comparison of observed values and predicted values (with 95% credible intervals) are depicted in Fig. 8. We report similar statistics for in-sample fits. Examples of in-sample fits for a selection of states are depicted in Fig. 9.

**Figure 8:**
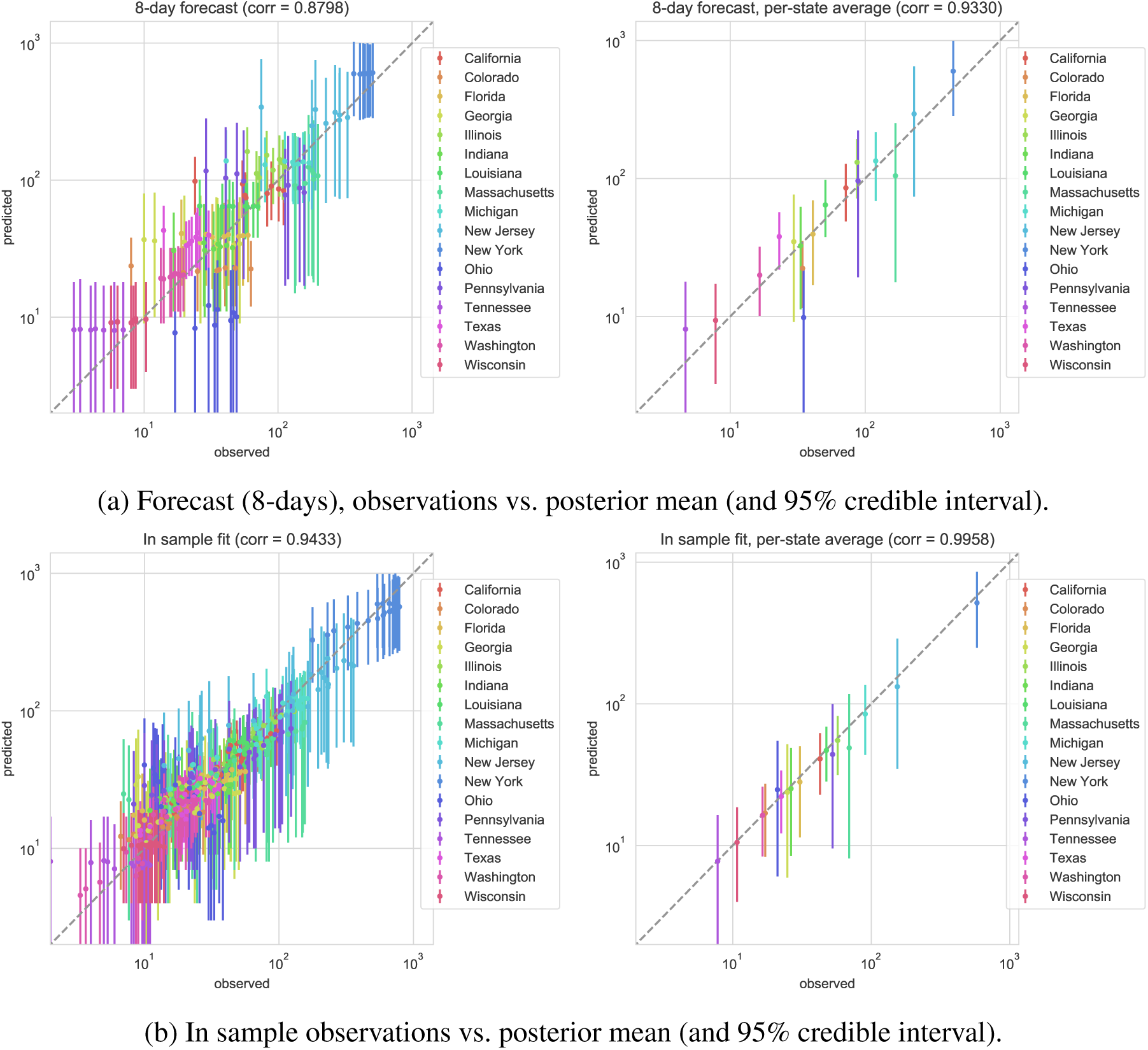
Eight day forecast (top) and in sample (bottom) model fits, comparing observed daily deaths (x-axis) and model posterior mean and 95% credible interval (y-axis). The left depicts each daily observation, the right depicts the average for each state.

**Figure 9:**
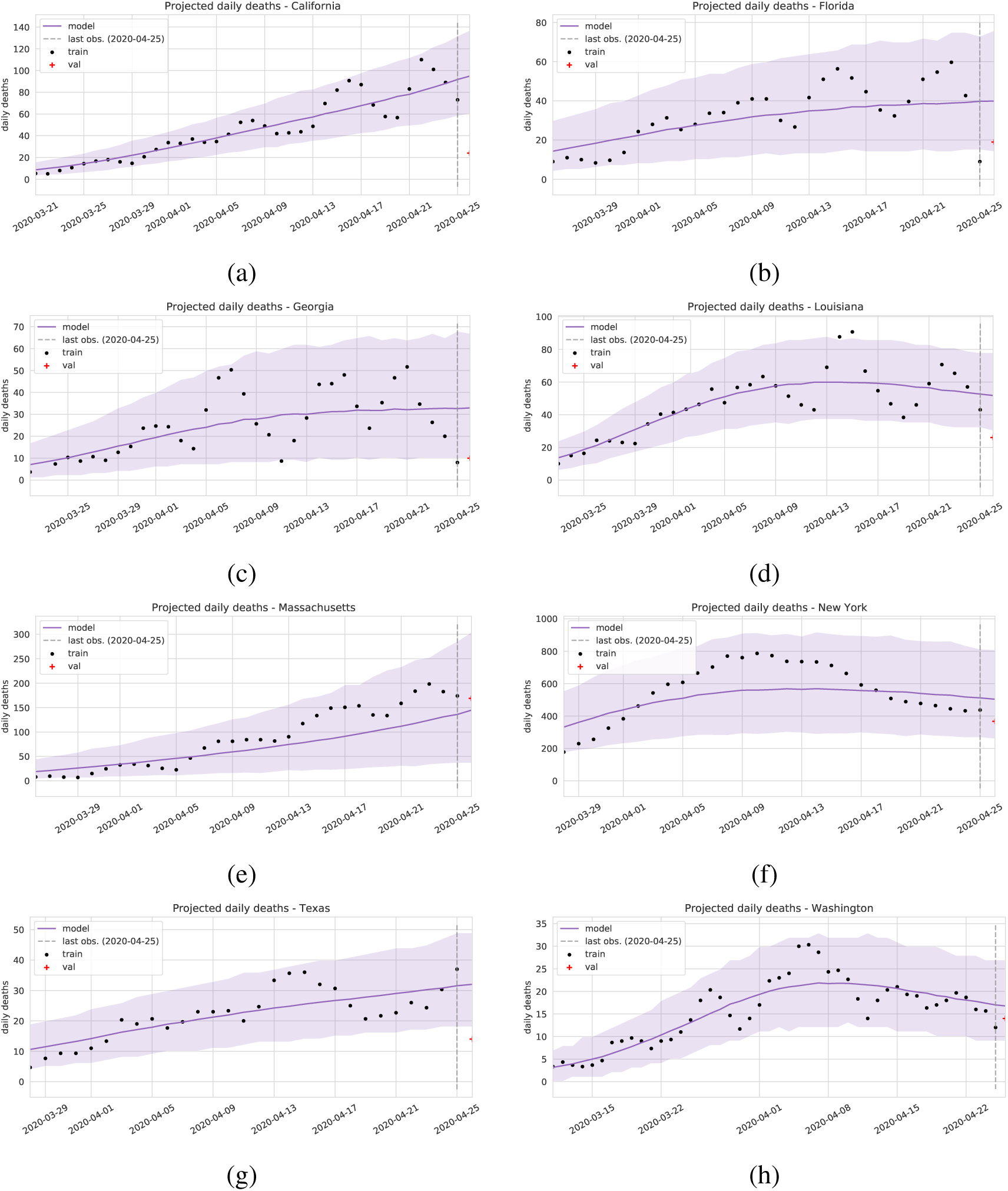
Model fit for daily deaths, mean and 95% credible interval. For some states, observations clearly exhibit a “day of week” effect. Florida, Georgia, New York, and Washington have shown more leveling off, whereas California, Massachusetts, and Texas appear to still be growing (as of April 25).

**Figure 10:**
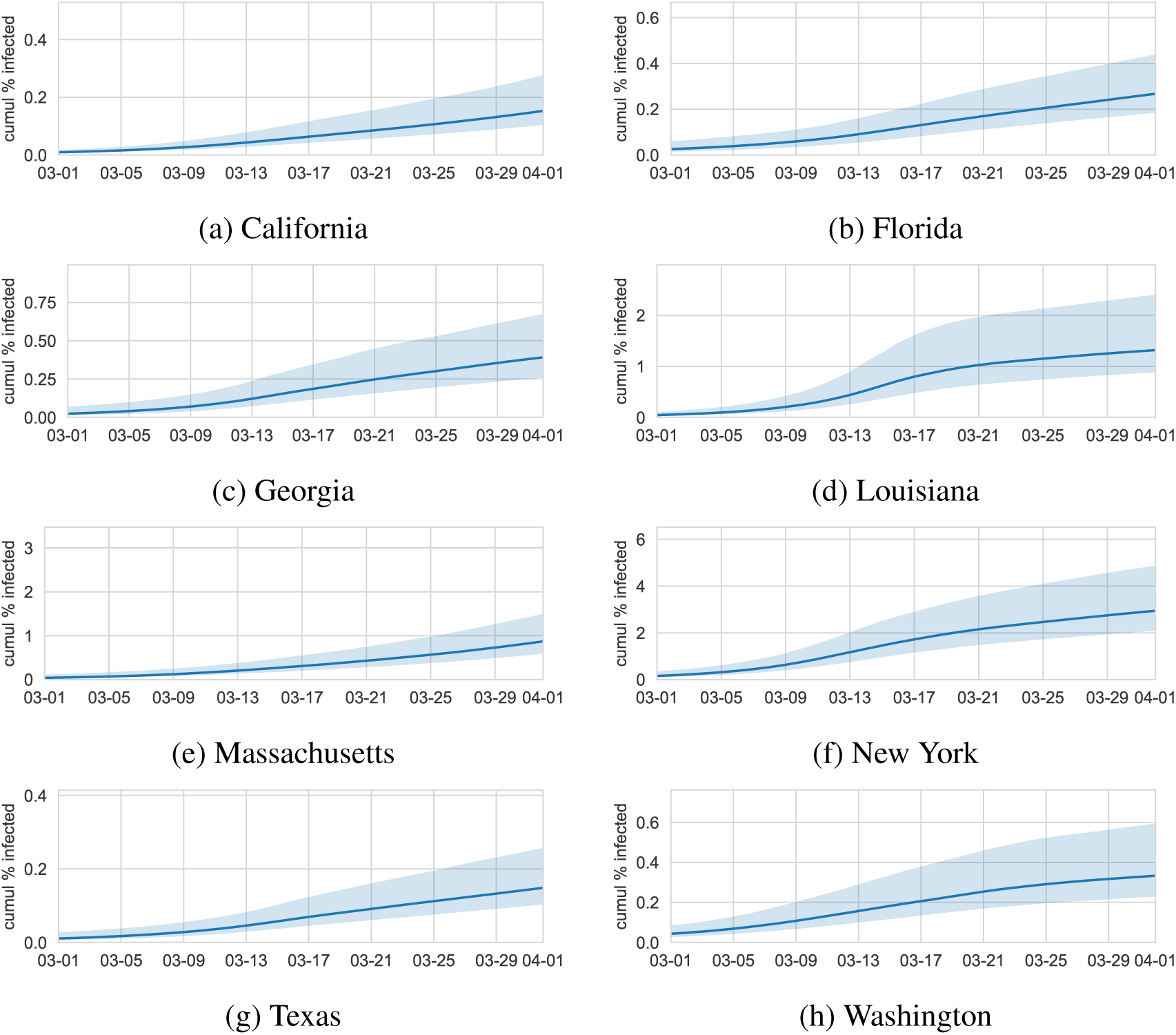
Percent of the population infected over time (cumulative) for eight states, posterior mean and 95% credible intervals shown.

**Figure 11:**
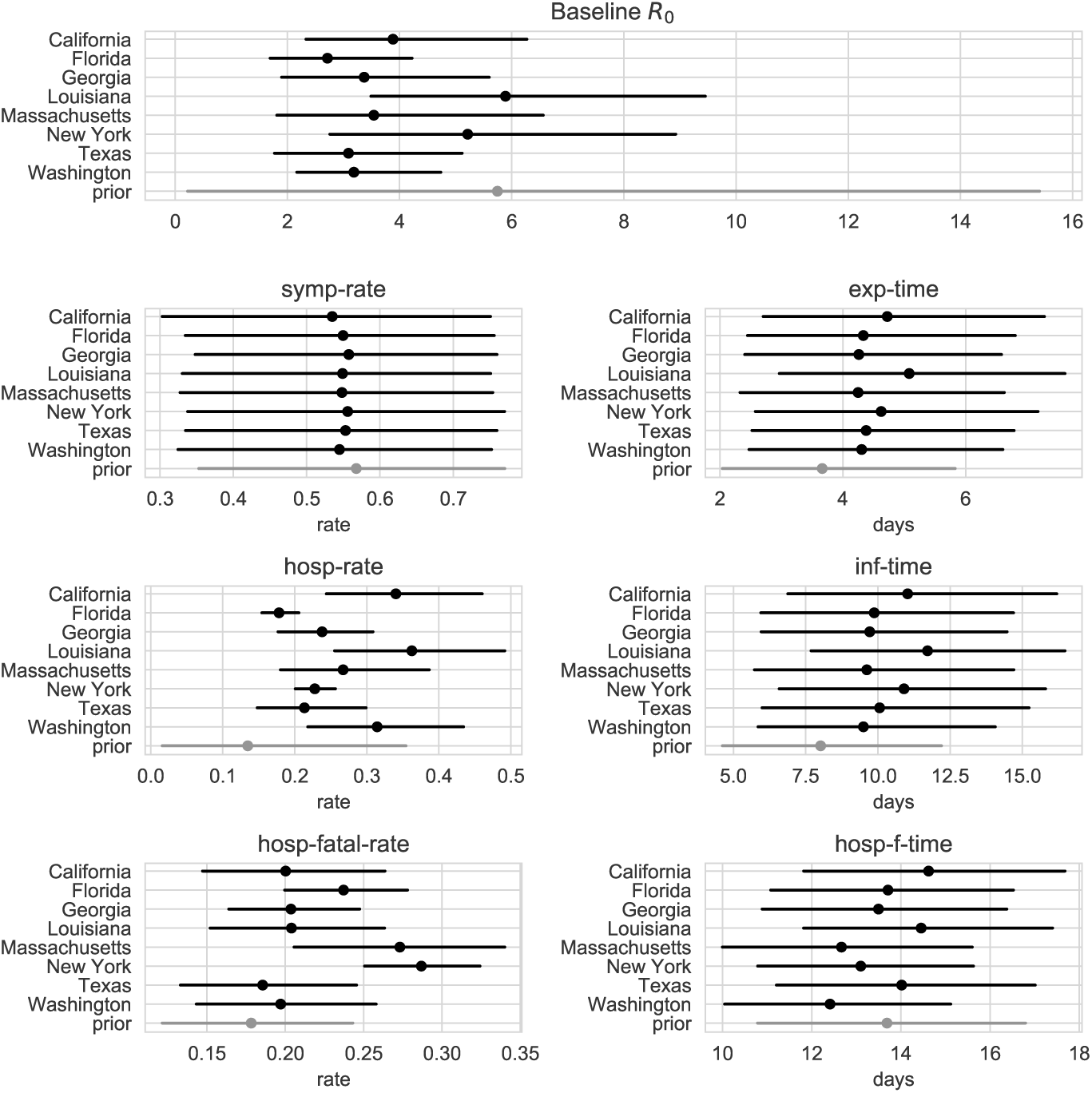
Inferred posterior times and rates between states, compared to the prior (in grey). Points are posterior means and lines span a 95% credible interval. Depicted is the ‘medium‘ strength prior over baseline *R*_0_.

### Sensitivity to prior strength

We explore the effect of the strength of our prior on baseline *R*_0_ for each state. As described above, we use three different strengths of prior on *R*_0_ with mean 2.4. For each prior strength, we visualize the relationship between *R*_0_*,_t_* and mobility volume, Mobility_t_, in Fig. 12. We find that the relationship between mobility volume and the *R*_0_ multiplier remains largely unchanged under the different prior strength, implying that the effect of mobility on transmission can be separated from other factors that affect the overall level of transmission such as population density.

**Figure 12:**
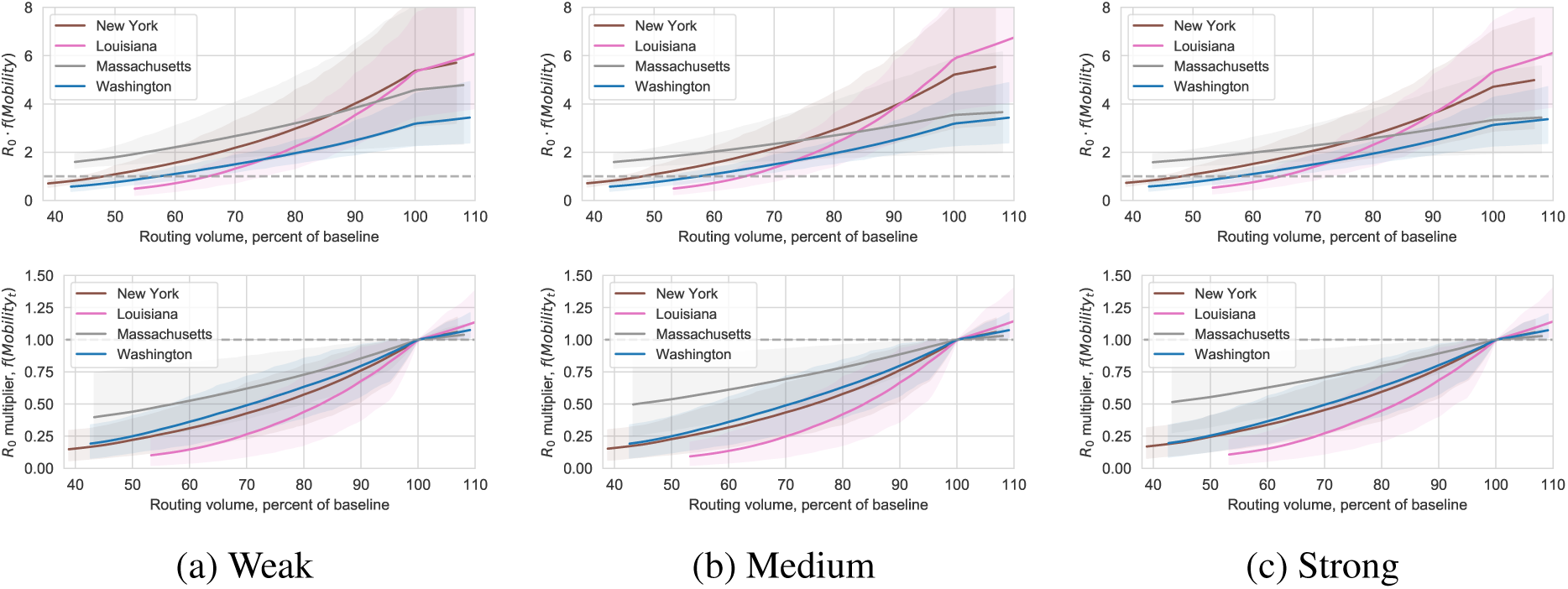
Sensitivity to R0 prior strength. The top panel shows the *R*_0_*_,t_* = *R*_0_ · *f*(Mobility*t*) value as a function of Mobility*t*; the bottom panel shows the baseline multiplier function *f*(Mobility*_t_*) (visualized with 95% posterior credible intervals). Left to right compares *R*_0_ prior strength (described in the SM) from least informative to most informative. While we see some sensitivity to the baseline *R*_0_ inferred (at 100% of routing baseline), credible intervals overlap considerably. The multiplier relationship between mobility and *f*(·) is stable across the three prior strengths.

### Accounting for importation

We extended our augmented SEIR model to include disease importation to account for elevated hospitalizations and deaths early in the epidemic. Specifically, we incorporated a constant importation parameter, *β*_imp_, into the force of infection. This resulted in the following dynamics equation for the *S* compartment:

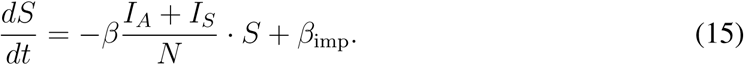

We placed a half-normal prior on with mean zero and variance one which constrains it to be greater than zero but allows the data to move it away from zero. We found that all of the states we consider the posterior over *β*_imp_ concentrates near zero and is much smaller than the initial fraction of exposed individuals. As such, we presented results in the paper using a version of the model without *β*_imp_. We account for these early effects by removing early observations, and fitting a distribution over the initial compartment values.

## Alternative Estimation of Effective R and Mobility Relationship

As further validation for the relationship between *R_E_,_t_* and routing volume inferred in the main paper, we recover a qualitatively similar relationship using confirmed case data and an independent estimation method. These results provide evidence that this relationship manifests in the data and does not arise only from the structure encoded in our model.

For this analysis we consider confirmed case counts for California, Louisiana, New York, and Washington. For each state we consider the time span from March 2nd to April 1st, except for Louisiana where the earliest confirmed non-zero case count is on March 9th. Unfortunately, there are issues with the confirmed case data due to limitations of testing. However, the case data exhibit a shorter lag from infection than the deaths data used for the model in the main paper and so they provide a good comparison data source.

We use the likelihood-based approach of Wallinga and Teunis (W&T) that estimates *R_E_,_t_* from the numbers of reported case counts (*25*). A key quantity needed for the W&T method is the *serial-interval distribution* that describes the time from symptom onset in a primary case to symptom onset in a secondary distribution. The R0 package for the R programming language was used to estimate *R_E_,_t_* (*33*). In Figs. 13–16 we depict the estimated *R_E_,_t_* over time using a Weibull serial-interval distribution with shape 5.4 and scale 4 (*17*). Note that there is a steady decrease in *R_E_,_t_* over time according to the case data. In each of these figures we also plot the estimated *R_E_,_t_* against the percent-change in routing volume. In each state we observe a decrease in *R_E,t_* corresponding to a decrease in routing volume. To quantify this relationship we used robust linear regression for the model *R_E_,_t_* = *β3Z_t_*+*b*. We fit the model to the simulated data described above. The resulting best fit lines are shown in Figs. 13–16 and are reported in Table 1. The coefficients are small but are statistically significant. In Fig. 17 we depict the inferred relationship for three serial-interval distributions with shape parameters 3, 5.4, and 8, (all with scale parameter 4). The relationship between *R_E_,_t_* and routing volume varies with the shape of the serial-interval distribution, growing as the shape parameter increases, similarly to the relationship learned in the main paper. Additionally, the variation differs between each shape indicating a dependence on the overall level of *R_E_,_t_*.

**Figure 13:**
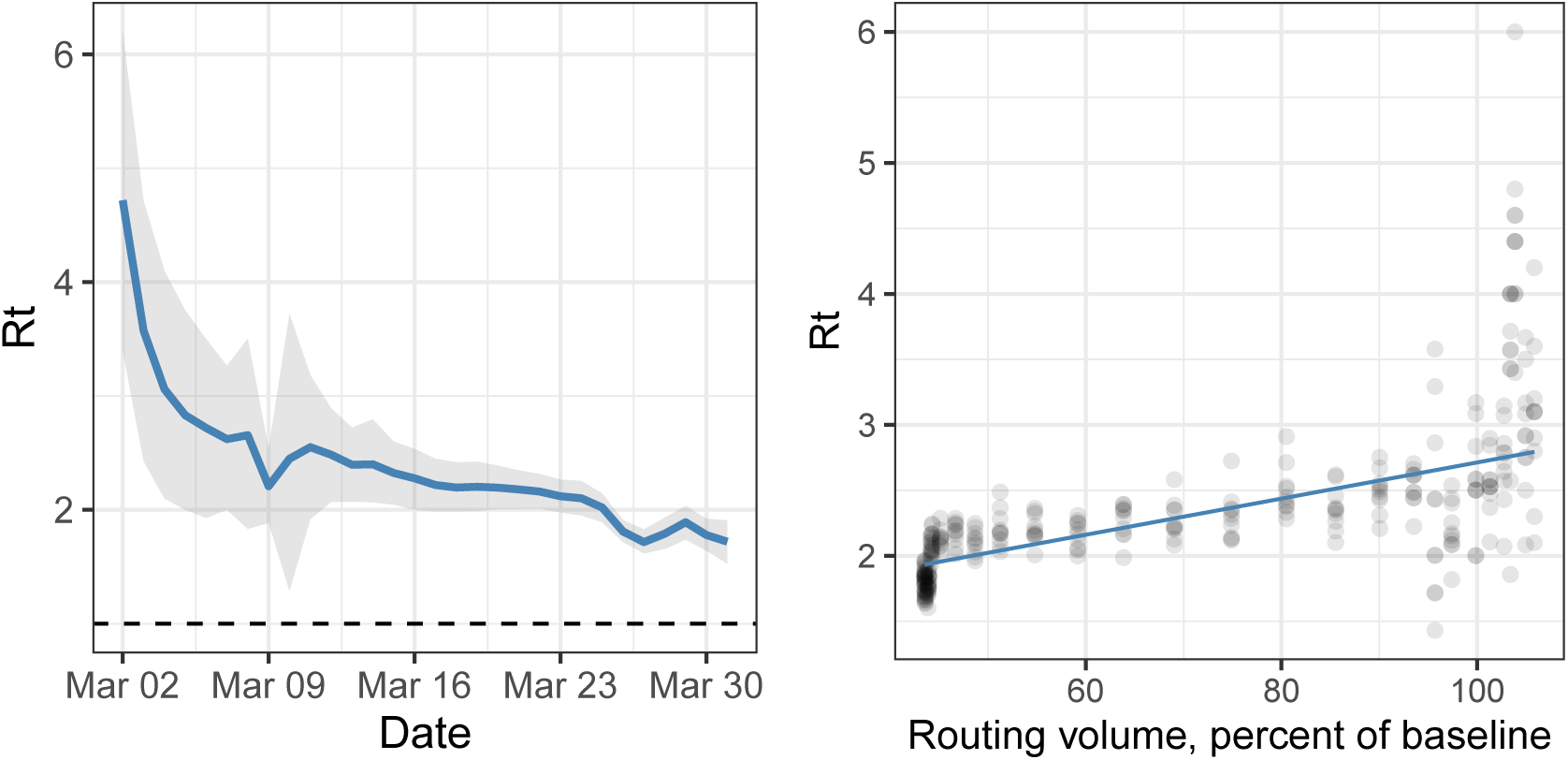
California: Estimated *R_E,t_* using the Wallinga and Teunis method and the resulting relationship between *R_E,t_* and percent-change in routing volume.

**Figure 14:**
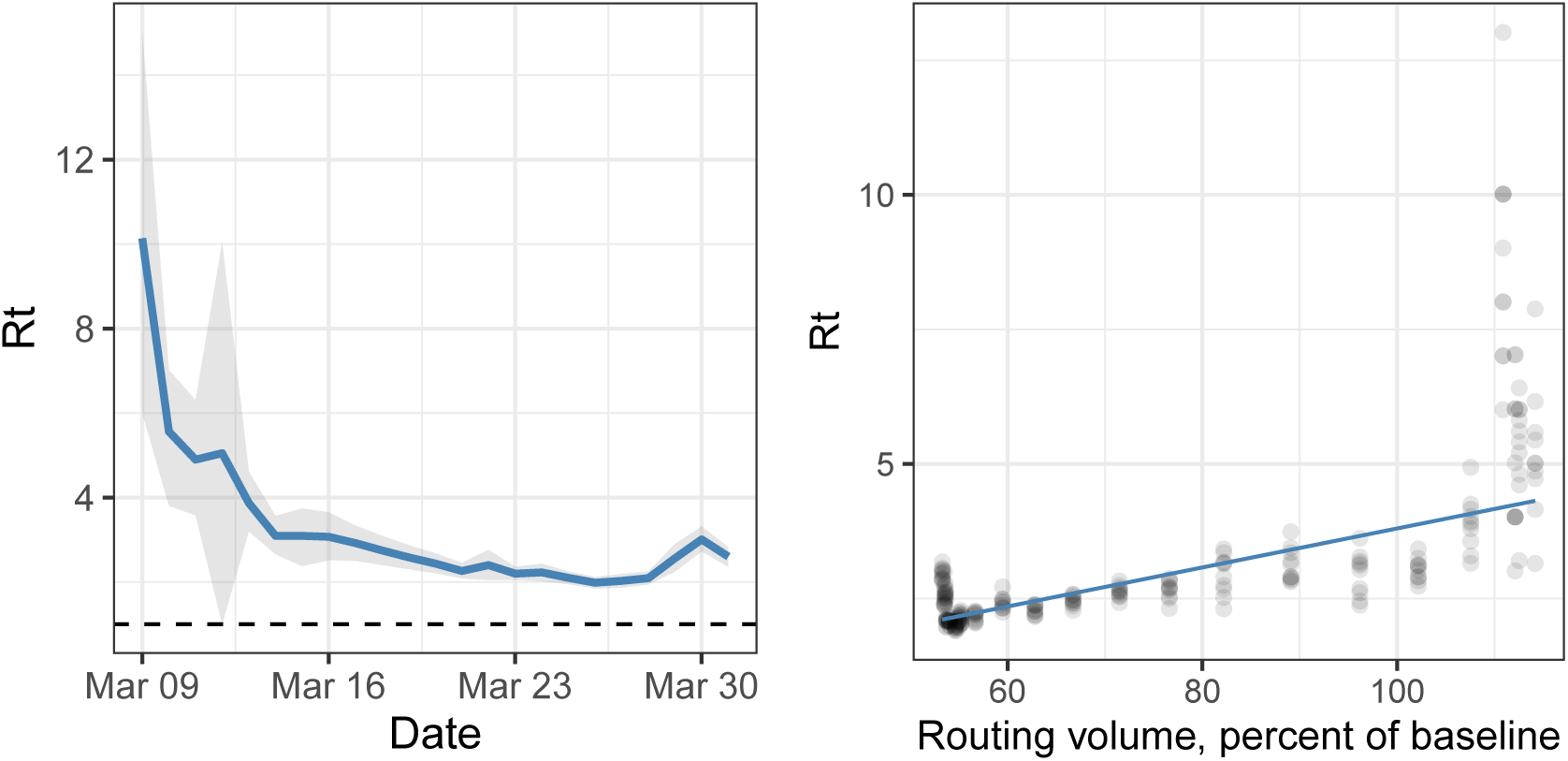
Louisiana: Estimated *R_E,t_* using the Wallinga and Teunis method and the resulting relationship between *R_E,t_* and percent-change in routing volume.

**Figure 15:**
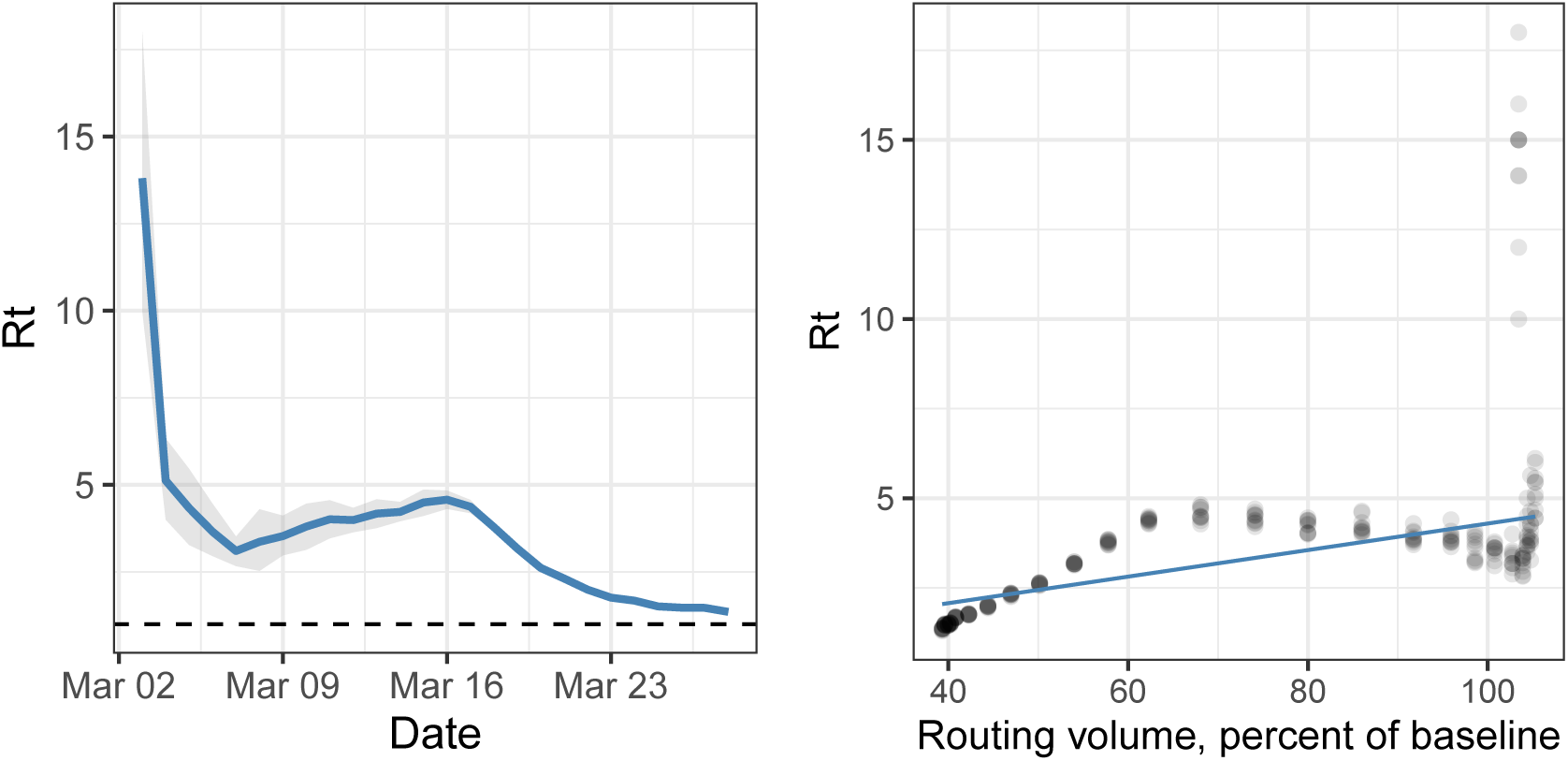
New York: Estimated *R_E,t_* using the Wallinga and Teunis method and the resulting relationship between *R_E,t_* and percent-change in routing volume.

**Figure 16:**
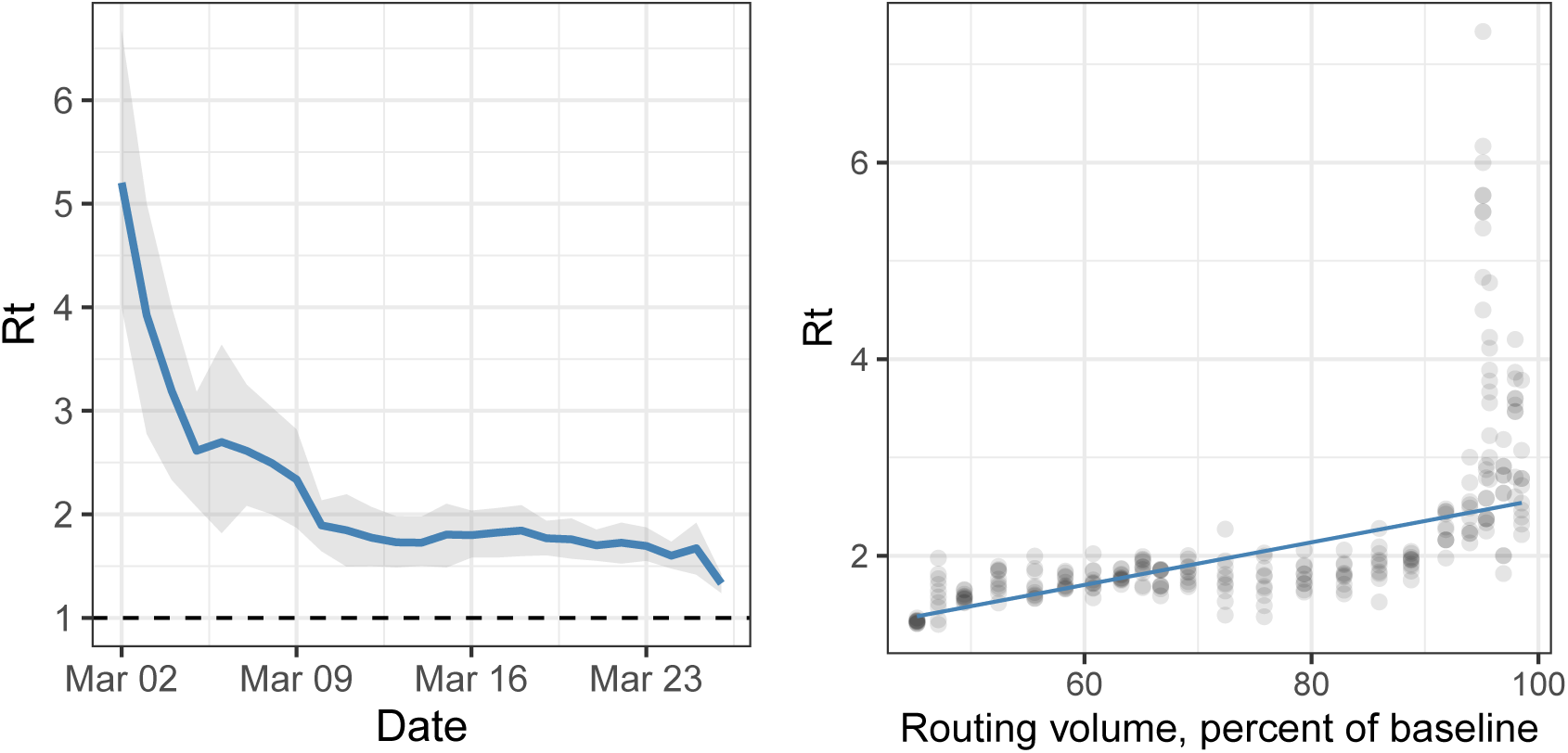
Washington: Estimated *R_E,t_* using theWallinga and Teunis method and the resulting relationship between *R_E,t_* and percent-change in routing volume.

**Figure 17:**
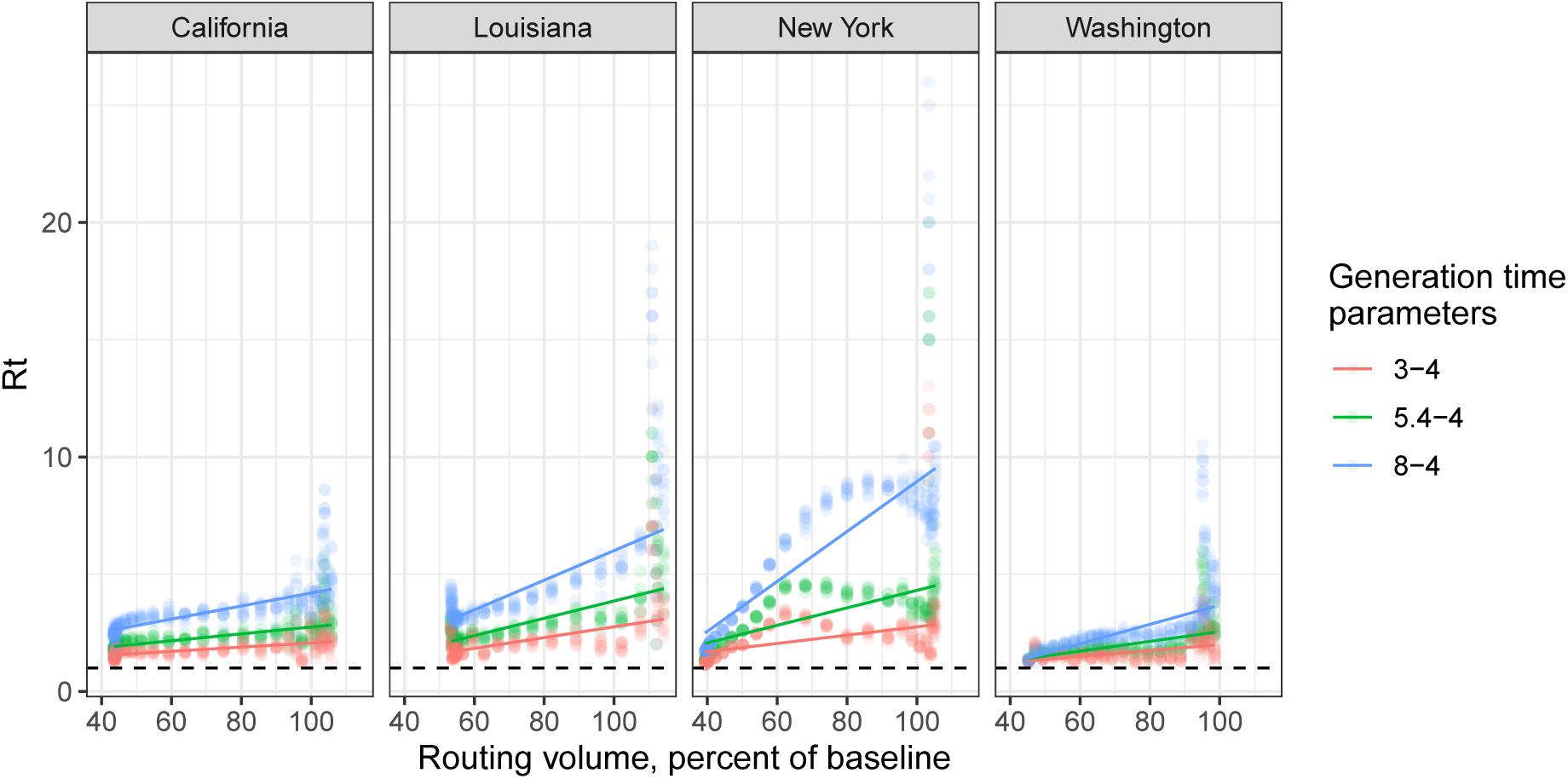
Comparison of *R_E,t_* vs. routing volume for each state using the Wallinga and Teunis method. Three serial-interval distributions with means 3, 5.4, and 8 days and variance 4. are depicted.

**Table 1:** Robust least squares estimates for a relationship between *R_E,t_* and routing volume.

| State | $\hat{\beta} \pm 1.96\text{s.e.}$ |
| --- | --- |
| California | $0.014 \pm 0.001$ |
| Louisiana | $0.036 \pm 0.002$ |
| New York | $0.037 \pm 0.004$ |
| Washington | $0.022 \pm 0.002$ |

The fact that the W&T method recovers a qualitatively similar relationship between mobility and *R_E_,_t_* provides extra evidence that our model is capturing an underlying relationship between mobility and disease transmission. Recall that the W&T model uses a disjoint set of data to estimate *R_E,t_* than our model indicating that this relationship is in the data and not imposed by our model.

## Data

### Deaths and hospitalizations counts

We use the daily counts of deaths from The New York Times which are based on reports from state and local health agencies (*21*). We include daily death observations once the state cumulative total has reached at least 40. This threshold filters out data while testing ramps up in states and when deaths counts may be inflated. Additionally, early deaths may not have resulted from community spread so that filtering out early deaths doesn't make the model explain these. For similar reasons, for New York we only include observations after March 25th, after a significant ramping up in daily tests levels off. We also filter out days where no deaths were reported since the previous day.

When available, we also use the number of hospitalized individuals in a state from The COVID Tracking Project (*22*). Similarly to the deaths data, we remove the first 100 hospitalizations from the data and remove days where no new hospitalizations were reported.

For both deaths and hospitalizations, we treat the days where data has been removed as missing data which do not contribute to the likelihood of our model and thus do not influence parameter estimates. However, uncertainty is propagated through these days due to our Bayesian formulation.

### Apple Maps mobility data

The data we use to quantify mobility has been collected by the Apple Maps team and consists of the relative number of directions requests (routes) per U. S. state relative to January 13, 2020. Apple does not store a profile of individual movement or searches as all data sent from users' devices to the Maps service are associated with random, rotating identifiers. Additionally, Apple Maps does not have any demographic information about users and so we cannot make any statements about the representativeness of the data in the larger population (*13*).

The state-level routing data exhibits a few quirks that must be addressed before being used in our modeling approach. First, the values are sensitive to the routing requests made on January 13, 2020. To address this we define the *baseline mobility* for a state as the average relative routing volume from January 13th to February 13th. We divide the relative routing values by this per-state baseline value and subtract 1 so that baseline mobility corresponds to zero.

There are also substantial day-of-week effects in the data with large dips in mobility on Sundays and increased mobility throughout the week. We remove these effects by smoothing the baseline-normalized data with a LOESS smoother. We use a value of 10/*T* for the fraction of data used to estimate each smoothed mobility value, where T is the number of days used by the model and varies for each state.

## Social distancing relaxation policy evaluation

In this section we present further results from the policy evaluation analysis. Fig. 18 depicts the same results as Fig. 5 from the main paper but with scales that show the entire 95% predictive intervals and make clear the uncertainy about the potential scale of the (secondary) outbreak in different states.

**Figure 18:**
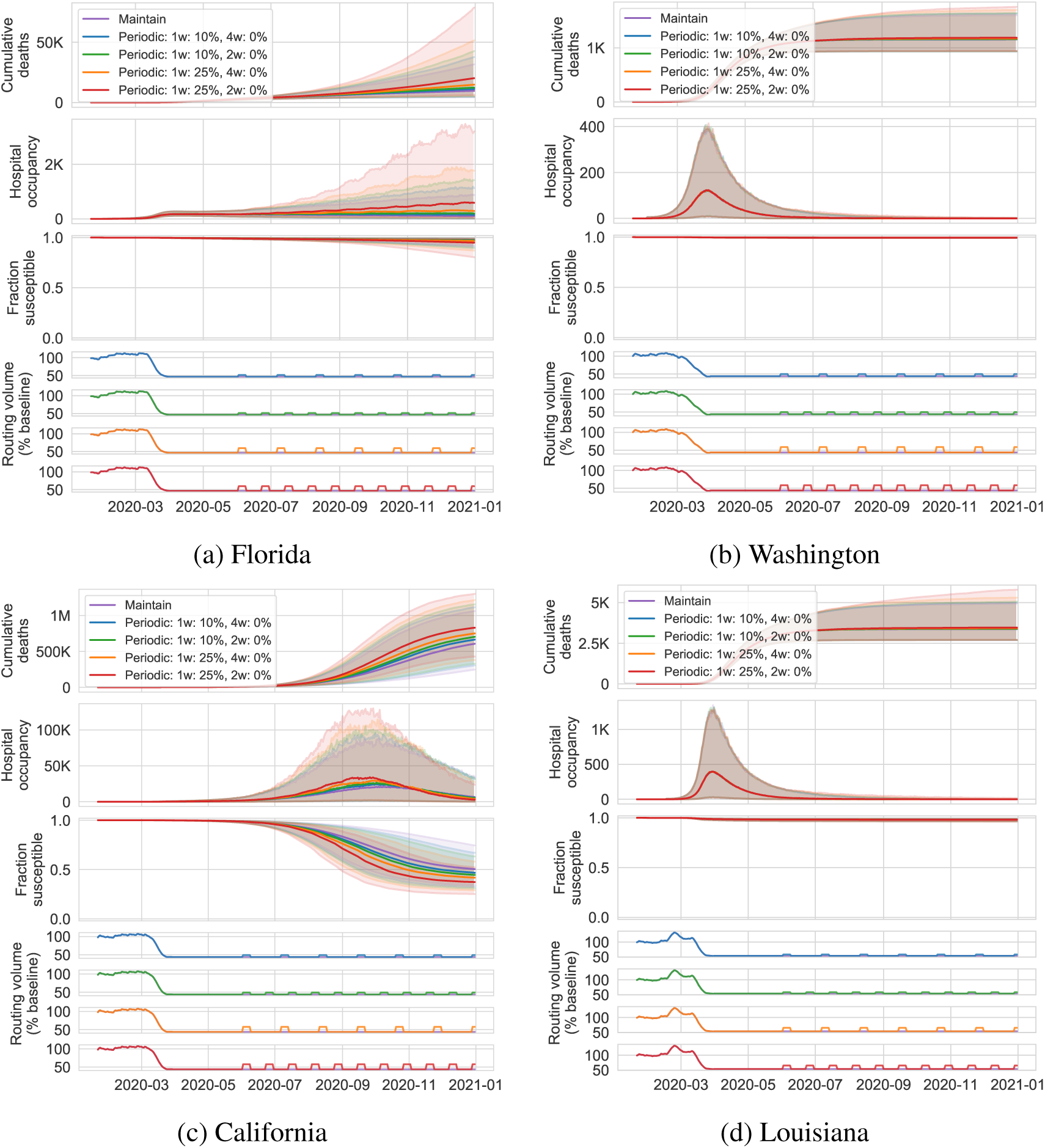
Median projections with 95% predictive intervals of cumulative deaths, hospital occupancy, and fraction-susceptible through December 31st, 2020 under the *periodic policy* for the states depicted in the paper. Axes scaled to include the entire 95% predictive intervals demonstrating the uncertainty in the scale of the outbreak.

Next, we present the predictions for Florida, Georgia, Texas, and Massachusetts for the lift policy (Fig. 19) and the periodic policy (Figs. 20 and 21). We see more variety in the shapes of the secondary outbreaks that depends on the initial *R*_0_ and the state-specific relationship between effective reproduction number and mobility.

**Figure 19:**
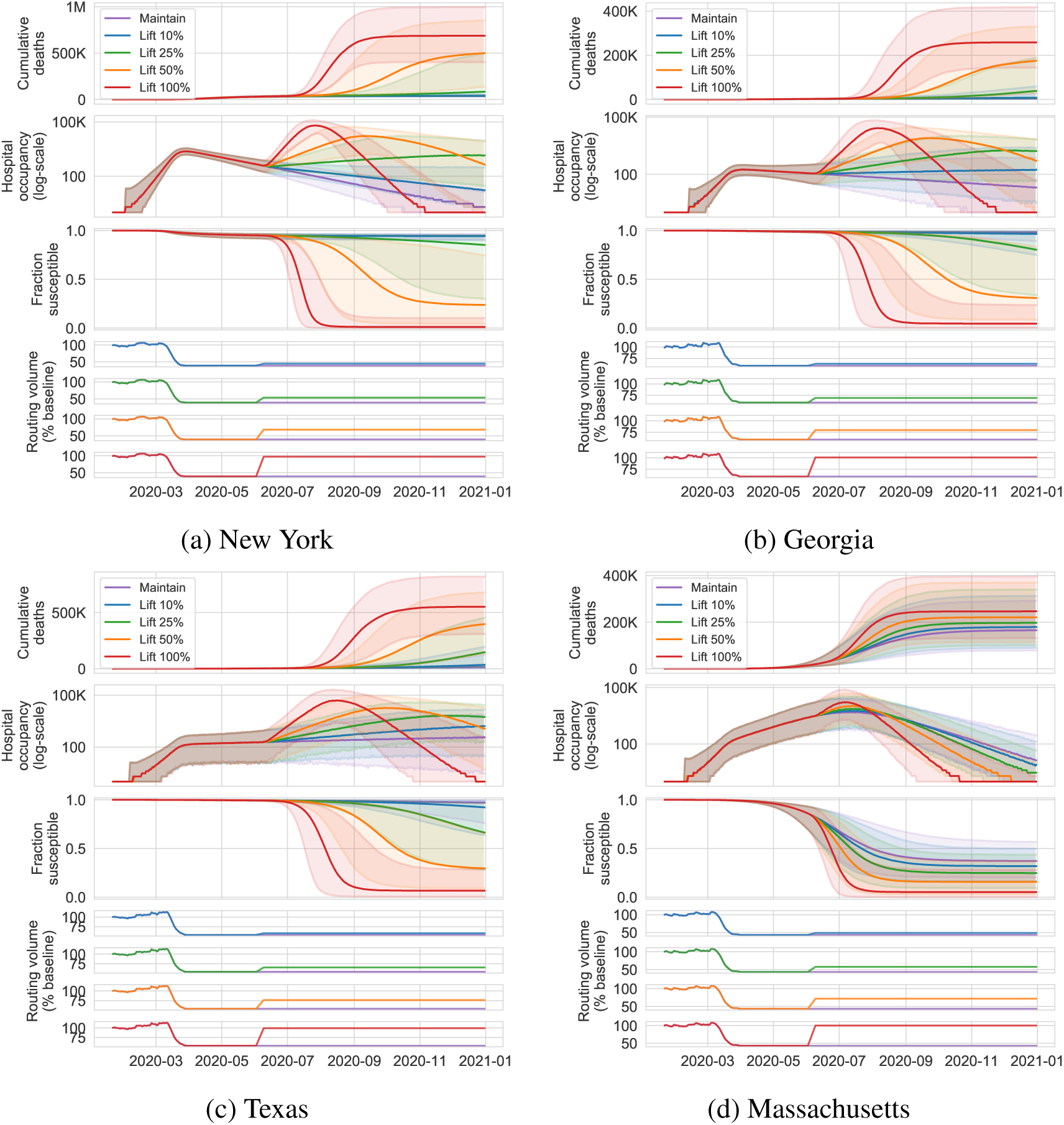
Median projections with 95% predictive intervals of cumulative deaths, hospital occupancy (log-scale), and fraction-susceptible through December 31st, 2020 under the *lift policy* for Florida, Georgia, Texas, and Massachusetts.

**Figure 20:**
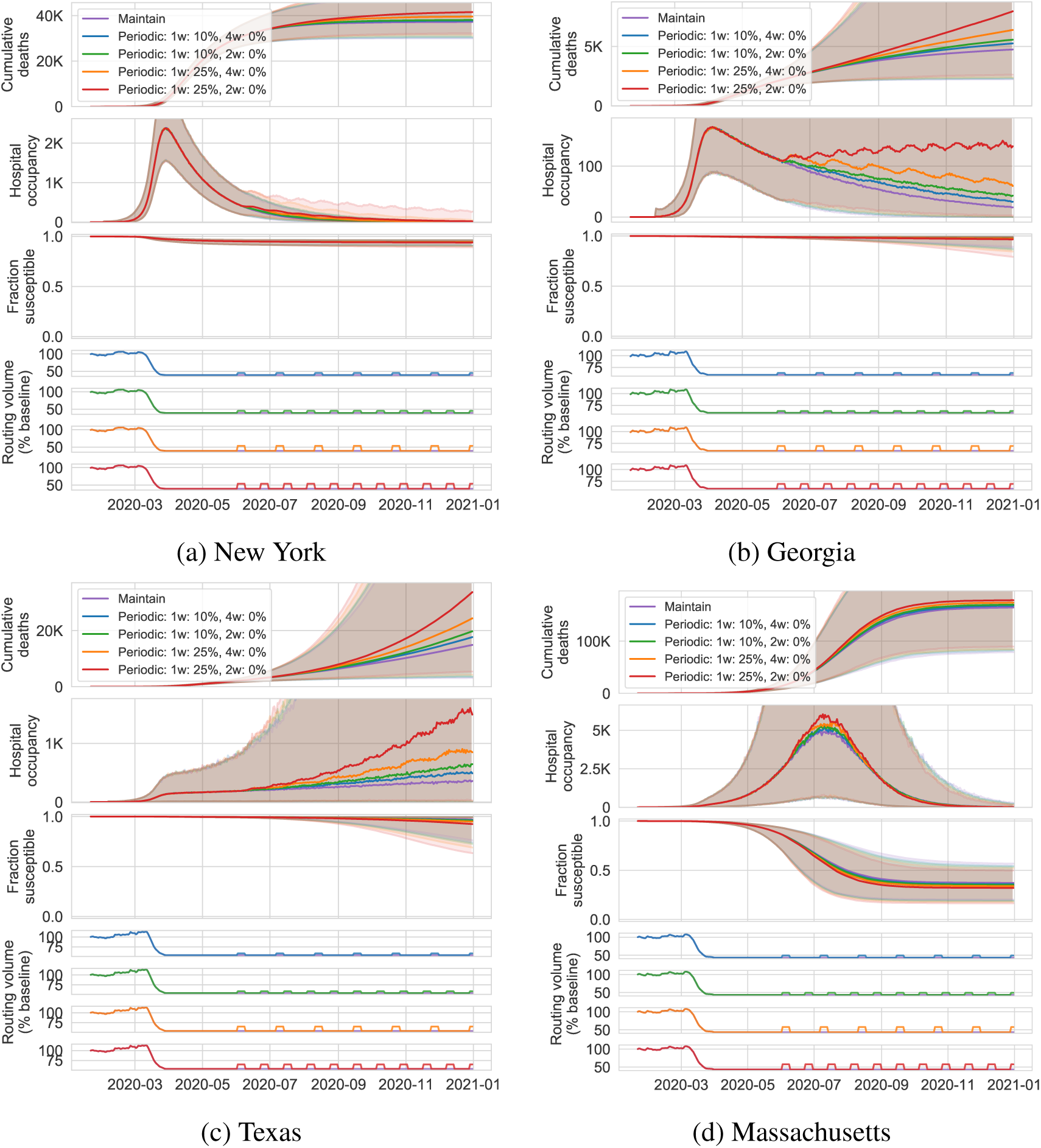
Median projections with 95% predictive intervals of cumulative deaths, hospital occupancy, and fraction-susceptible through December 31st, 2020 under the *periodic policy* for New York, Georgia, Texas, and Massachusetts. Plots zoomed in on the median projections.

**Figure 21:**
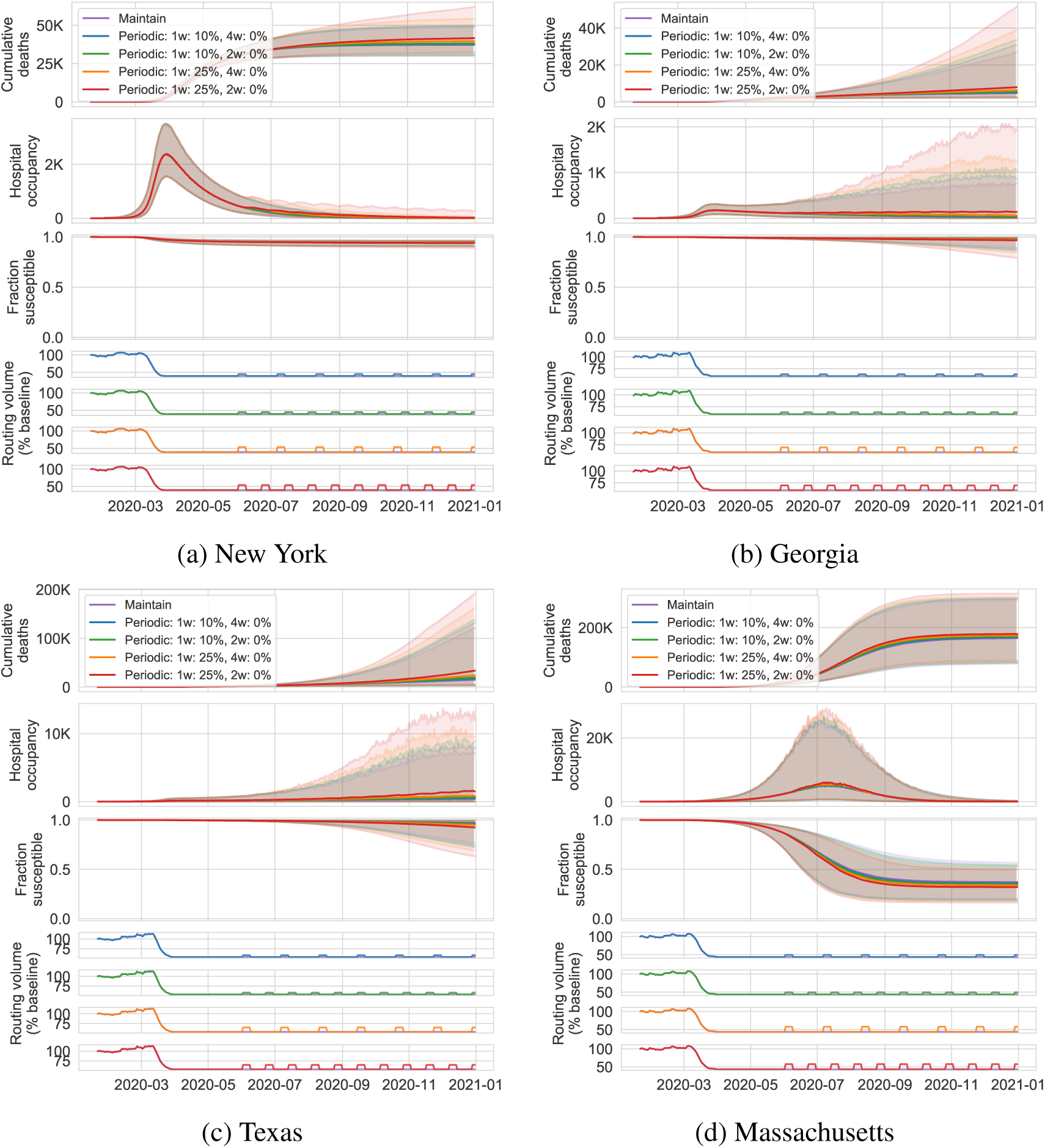
Median projections with 95% predictive intervals of cumulative deaths, hospital occupancy, and fraction-susceptible through December 31st, 2020 under the *periodic policy* for New York, Georgia, Texas, and Massachusetts.

